# HOW SAFE IS EXERCISE? CROSS-SECTIONAL ASSOCIATIONS OF CURRENT PHYSICAL ACTIVITY WITH SELF-REPORTED FRACTURE HISTORY IN MID-LIFE

**DOI:** 10.64898/2026.07.27.26358989

**Authors:** Catherine Rolls, Jonathan H Tobias, Emma M Clark, Helen Dawes, Marcus Munafo, Benjamin G. Faber

**Author notes:** **Corresponding author – Catherine Rolls**.

## Abstract

**Background:** Physical activity is important for healthy ageing, but the relationship between the volume and intensity of activity and fracture during mid-life remains unclear, particularly at high activity levels. Understanding whether higher volumes or intensities of activity are associated with fracture is important when promoting physical activity during mid-life. We examined associations between the volume and intensity of leisure-time physical activity and fracture, and whether adherence to World Health Organization (WHO) physical activity recommendations was associated with fracture.

**Methods:** Cross-sectional analysis of UK Biobank participants aged 40–65 years using self-reported physical activity and fracture data. Leisure-time physical activity volume (LTPAv) was derived from walking, moderate and vigorous activity and expressed as metabolic equivalent task (MET)–minutes/week. Participants were categorised into LTPA_V_ quintiles and by adherence to WHO physical activity recommendations. Multivariable logistic regression examined associations with self-reported fracture within the previous five years, with analyses adjusted for age, ethnicity, education, deprivation, weight and height, and stratified by sex. To understand the independent effects of intensity: walking, moderate and vigorous activity volume (MET-minutes/week) were included simultaneously in mutually adjusted models.

**Results:** Among 322,749 participants, 53% were female with a mean age of 55 years. Odds of fracture increased with higher LTPA_V_, although the association was non-linear and varied by age and sex. The strongest associations were in the highest quintile among men and women aged 40–49 years (OR 1.81, 95% CI 1.64–1.99 and OR 1.42, 95% CI 1.28–1.58 respectively). In mutually adjusted models, vigorous activity demonstrated the strongest independent association with odds of fracture. Meeting WHO physical activity recommendations, without exceeding them, was not associated with increased odds of fracture.

**Conclusions:** Higher volumes of leisure-time physical activity were associated with greater odds of fracture, with associations largely driven by vigorous-intensity activity and strongest in adults aged 40–49 years and in men. Activity consistent with current WHO recommendations was not associated with greater odds of fracture. These findings support continued promotion of physical activity during mid-life while highlighting the need to better understand fracture risk among individuals undertaking high volumes of vigorous activity.

## BACKGROUND

Fractures during mid-life (40–65 years) are associated with substantial morbidity, healthcare utilisation and economic burden. Among working-age adults, extremity fractures have been linked to more than 1,700 hours of lost work and costs exceeding US$64,000 per patient during the year after injury.^1–3^ Fracture incidence differs markedly by sex with men experiencing higher fracture rates during adolescence and early adulthood, whereas fracture incidence in women rises rapidly between 50 and 60 years of age, coinciding with the menopause.^4–6^ Menopause is also associated with changes in bone microarchitecture, muscle mass and physical function that may further increase fracture risk.^7,8^ Consequently, this represents a potentially important period for preventive strategies, including physical activity, aimed at maintaining musculoskeletal health and reducing future fracture risk.

Leisure-time physical activity (LTPA) during mid-life is associated with lower multimorbidity and healthier ageing,^9,10^ with even an increase in light activity being found to reduce mortality for those less able to exercise at high levels.^11^ The World Health Organisation (WHO) recommends that adults undertake 150–300 minutes of moderate-intensity, or 75–150 minutes of vigorous-intensity physical activity weekly, the upper threshold representing an optimal target intended to achieve additional health benefits. These recommendations are primarily based on evidence relating to improved cardiovascular, metabolic and cancer outcomes,^12–14^ however, possible adverse health effects from physical activity levels exceeding the upper target are currently unclear. The relationship between LTPA and fracture risk is likely to be more complex than for other health outcomes. While LTPA is expected to improve musculoskeletal health through beneficial effects on bone and muscle strength, higher-intensity or higher-volume activity may also increase exposure to falls, trauma and sports-related injury.^4,15^ Consequently, LTPA may act as both a protective and risk factor for fracture depending on activity intensity, type and volume. Understanding this balance may be particularly important during mid-life, when maintaining physical activity has important implications for long-term health, yet concerns regarding injury and fracture risk may influence participation.

Physical activity levels decline during mid-life, particularly among women. Qualitative studies suggest that women may reduce or modify LTPA around the menopause because of concerns about injury, falls and fracture risk.^16,17^ Despite these concerns, evidence-based recommendations regarding safe levels and intensities of LTPA in relation to fracture risk during mid-life remain limited, and current menopause and osteoporosis guidelines provide only broad recommendations regarding physical activity.^18,19^ Relatively few studies have focused specifically on mid-life populations. A recent systematic review identified substantial uncertainty regarding both the direction and magnitude of the association between physical activity and fracture risk in mid-life,^20^ with much of the available evidence relying on surrogate outcomes such as bone mineral density, rather than clinically relevant fracture endpoints.^7,21,22^ In particular, the relative contributions of activity volume and intensity to fracture risk, and whether these associations differ by age and sex, remain unclear. Therefore, the aim of this study was to determine whether LTPA volume and intensity are associated with fracture risk during mid-life and whether these associations differ by age and sex, using data from the UK Biobank (UKB).

## MATERIALS AND METHODS

### Study design and population

This study is an observational cross-sectional analysis of self-reported LTPA and self- reported fracture in the UKB, a prospective cohort study of over 500,000 adults aged 40–69 years recruited throughout the UK between 2006 and 2010. Details of recruitment and data collection have been described previously.^23^ This research was conducted using UKB resources under application number 81499.

### Physical activity

For self-reported LTPA, we used data collected at the baseline assessment centre visit (2006– 2010). UKB participants completed questions derived from the validated International Physical Activity Questionnaire,^24^ which assessed leisure-time walking, moderate-intensity and vigorous-intensity physical activity undertaken during the previous week. Frequency (days/week) and duration (minutes/day) of each activity were used to calculate weekly volumes of walking, moderate and vigorous LTPA, expressed as minutes/week. Metabolic equivalent task (MET)-minutes/week were then calculated for each activity by multiplying weekly volume by the corresponding established intensity weights.^24^ A composite LTPA volume measure (LTPA_V_) was derived by summing the activity-specific MET-minutes/week values.

### Fracture ascertainment

Our primary outcome was self-reported fracture within the previous five years, collected at the baseline assessment. As fractures sustained during mid-life may not require hospital admission, self-reported fracture was selected to capture fractures that may not be identified through hospital records. This was operationalised as ‘any fracture’. Participants reporting a fracture were prompted to identify the fracture site from a pre-specified list comprising hip, spine, wrist, arm, leg, ankle and other. Fracture-site information was used descriptively to characterise the distribution of reported fractures within the cohort.

### Covariates

Confounders were identified a priori using a directed acyclic graph (Supplementary Figure S1), from which a minimally sufficient adjustment set was derived. Age was participants’ age at the baseline assessment centre visit and stratified into 10-year age bands, (40-49, 50-59) and 60-65. The top age band capped at 65 as per the cohort age description. Ethnicity was self-reported and grouped as White, Asian and British Asian, Black and Black British, or Mixed ethnicity. Educational attainment was categorised as school education or less, college or vocational training, and university degree or above. Socioeconomic deprivation was measured using the Townsend Deprivation Index, with higher scores indicating greater material deprivation.^25^ Standing weight and height were measured at the baseline assessment centre visit according to UKB protocols.^26^

### Statistical analysis

The study is reported in line with the Strengthening the Reporting of Observational Studies in Epidemiology (STROBE) guidelines^27^, and the Checklist for statistical Assessment of Medical Papers (CHAMP)^28^. The study protocol was pre-registered,^29^ and minor deviations from the protocol are documented in Supplementary Methods S1. Code is available at GitHub (https://github.com/RollsCf/UKB_PA_Fracture.git).

Continuous variables are presented as mean (standard deviation (SD)) or median (interquartile range (IQR)), and categorical variables as frequency (percentage). Analyses were restricted to participants with complete data for the exposure, outcome and covariates. Baseline characteristics of included and excluded participants were compared using standardised mean differences.

The association between self-reported LTPA_V_ and self-reported fracture was examined using logistic regression. Models were constructed with increasing levels of adjustment: (1) unadjusted; (2) minimally adjusted for age (in years), sex, ethnicity, weight and height; and (3) fully adjusted, additionally including socioeconomic deprivation and education. Potential non-linear dose–response associations between total physical activity and odds of fracture were assessed using restricted cubic spline models within fully adjusted regression analyses. To aid interpretation, LTPA_V_ was modelled using quintiles of the raw exposure, with the lowest activity quintile (Q1) as the reference group. Additional analyses examined activity intensity-specific associations. To understand the independent effects of intensity, walking, moderate and vigorous activity volume (MET-minutes/week) were modelled separately, with each model mutually adjusted for the other two intensity-specific measures. Effect modification by age and sex was assessed through inclusion of interaction terms and subsequently stratified where appropriate. To contextualise findings relative to public health recommendations participants were additionally categorised according to whether they did not meet, met, or exceeded WHO physical activity recommendations. (Supplementary Methods S2).

Sensitivity analyses examined the proportion of total activity accumulated at each intensity level, alternative measures of body size (body mass index and waist circumference), and physical activity frequency (days/week) as an alternative to the primary MET-based exposure measure. All analyses were conducted using R (version 4.5.2).

## RESULTS

### Population characteristics

There were 322,749 individuals in the final study. Fifty four percent of the cohort were female, with a mean age of approximately 55 years (SD 7.3). Ninety-five percent of the cohort were of white ethnicity (Table 1) Missingness was low for most variables (<5%), although higher for physical activity measures, particularly LTPA_V_ (22.4%). Participants excluded because of missing exposure or covariate data were broadly similar to those included in the analysis, with no evidence of substantial imbalance in measured baseline characteristics (Supplementary Tables S1-2). Figure 1 shows flow of participants through the study.

**Figure 1:**
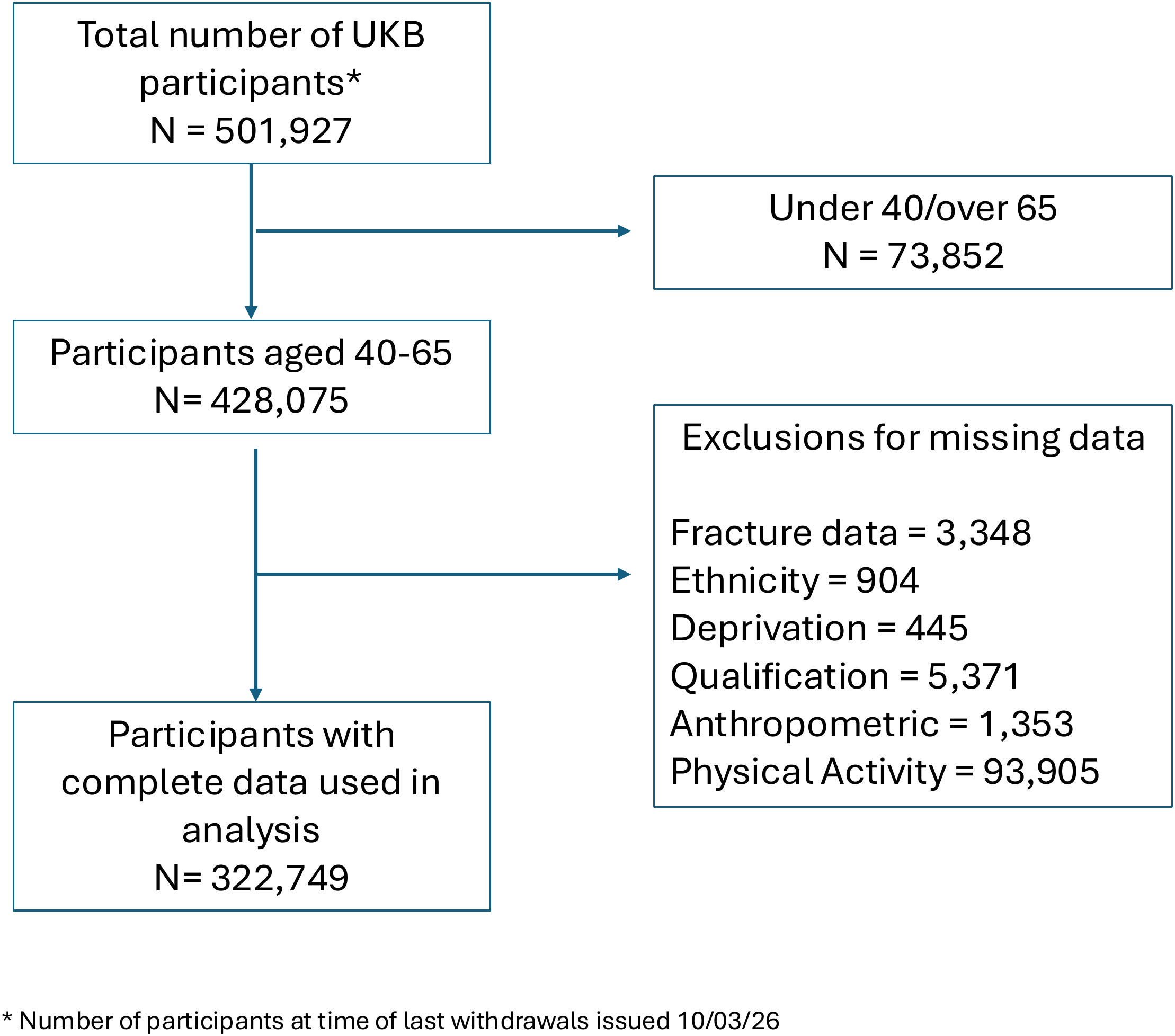
Flow diagram of participant selection for the study. Flow diagram showing the selection of UK Biobank participants included in the analyses of leisure-time physical activity (LTPA) and self-reported fracture. Participants were excluded sequentially according to the criteria shown. The final analytical cohort comprised participants aged 40–65 years with complete data for the exposure, outcome and covariates included in the analyses

**Table 1:** Baseline characteristics of participants by sex (n=322,749)

| **Variable** | **Sex** |  |  |
|---|---|---|---|
|  | **Overall** N = 322,749<sup>*1*</sup> | **Female** N = 171,040<sup>*1*</sup> | **Male** N = 151,709<sup>*1*</sup> |
| Age (years), mean (SD) | 54.5 (7.3) | 54.3 (7.2) | 54.7 (7.3) |
| Ethnicity, n (%) |  |  |  |
| Asian | 5,578 (1.7%) | 2,551 (1.5%) | 3,027 (2.0%) |
| Black or other | 8,724 (2.7%) | 4,994 (2.9%) | 3,730 (2.5%) |
| Mixed | 2,035 (0.6%) | 1,266 (0.7%) | 769 (0.5%) |
| White | 306,412 (95%) | 162,229 (95%) | 144,183 (95%) |
| Townsend deprivation index, mean (SD) | -1.4 (3.0) | -1.4 (3.0) | -1.4 (3.1) |
| Education level, n (%) |  |  |  |
| School education or less | 271,094 (84%) | 144,170 (84%) | 126,924 (84%) |
| College or vocational training | 30,766 (9.5%) | 15,462 (9.0%) | 15,304 (10%) |
| University degree or above | 20,889 (6.5%) | 11,408 (6.7%) | 9,481 (6.2%) |
| Weight (kg), mean (SD) | 78.3 (16.0) | 71.2 (14.0) | 86.2 (14.3) |
| Height (cm), mean (SD) | 169.2 (9.3) | 163.0 (6.3) | 176.2 (6.8) |
| BMI (kg/m<sup>2</sup>), mean (SD) | 27.2 (4.7) | 26.8 (5.1) | 27.8 (4.2) |
| Fracture in the last 5 years, n (%) |  |  |  |
| No | 292,661 (91%) | 154,478 (90%) | 138,183 (91%) |
| Yes | 30,088 (9.3%) | 16,562 (9.7%) | 13,526 (8.9%) |
| Moderate intensity activity volume (MET-min/week), median (IQR) | 480 (120, 1,200) | 480 (120, 1,200) | 420 (120, 1,200) |
| Vigorous intensity activity volume (MET-min/week), median (IQR) | 240 (0, 960) | 192 (0, 720) | 320 (0, 1,000) |
| Walking volume (MET-min/week), median (IQR) | 693 (297, 1,386) | 693 (330, 1,386) | 693 (297, 1,386) |
| Leisure-time physical activity volume (LTPAv) (MET-min/week), median (IQR) | 1,770 (813, 3,546) | 1,733 (810, 3,375) | 1,817 (813, 3,726) |

<sup>*1*</sup>Continuous demographic and anthropometric variables are presented as mean (SD). Physical activity variables are presented as median (IQR). Categorical variables are presented as n/N (%).

### Fracture distribution

Fracture within the previous five years was reported by 16,562 (9.7%) of women and 13,526 (8.9%) men, corresponding to 30,088 (9.3%) self-reported fractures across the cohort.

Fracture-site distributions were broadly similar across sex and age groups (Figure 2, Panel A). Most fractures were classified as “other” sites (39-61%), while wrist fractures represented the most common specified fracture type, particularly among women aged 60-65 years (25.1%). Hip fractures were uncommon (<3%) but increased modestly with age in both sexes. (Supplementary Tables S3-4)

**Figure 2:**
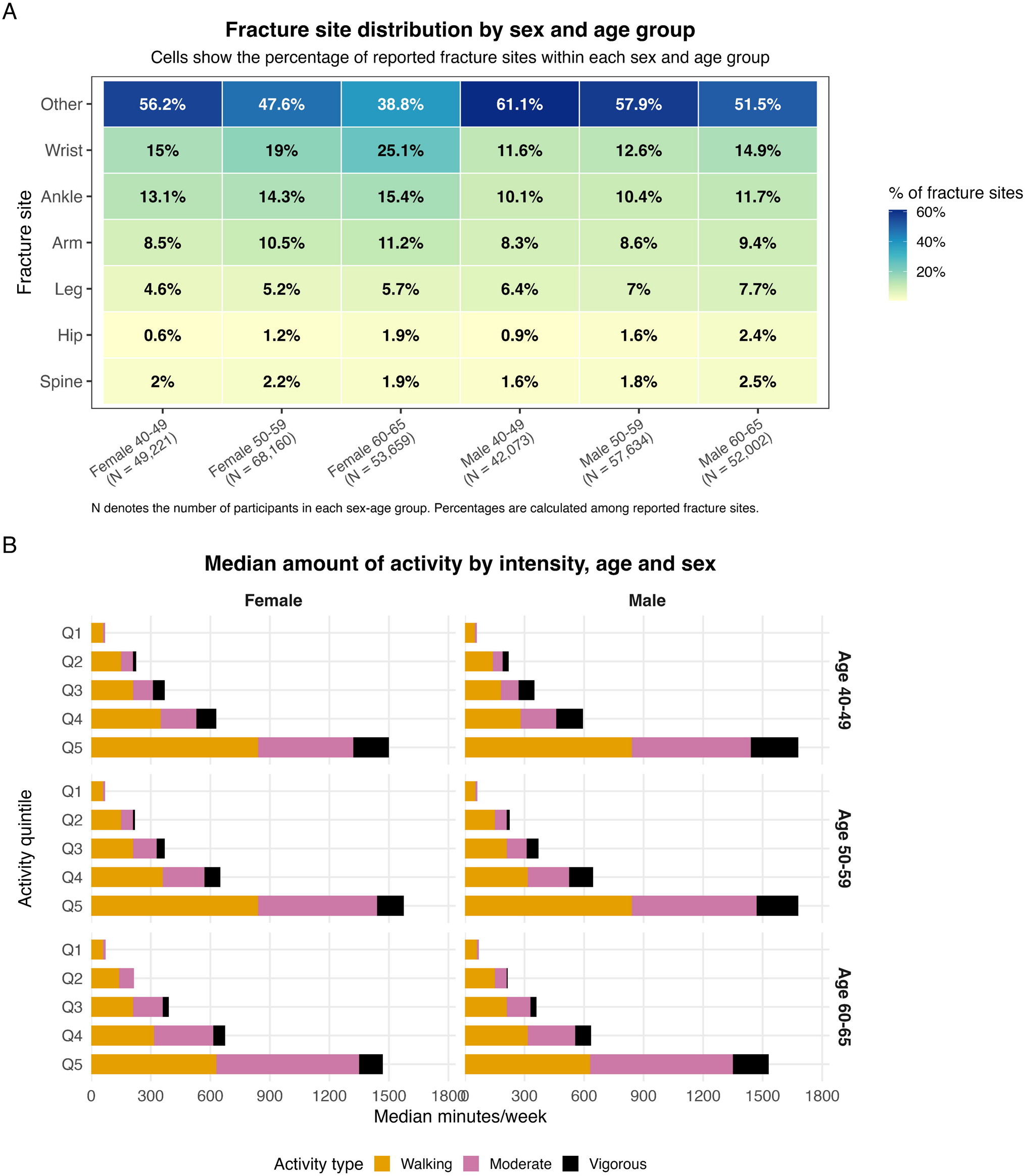
Fracture site distribution and activity intensity distribution by sex and age group. (Panel A) Distribution of self-reported fracture sites among participants reporting a fracture within the previous five years, presented as the percentage of fracture sites within each sex-age group. (Panel B) Median weekly minutes of walking, moderate and vigorous physical activity within each quintile of leisure-time physical activity volume, stratified by sex and age group.

### Distributions and characteristics of physical activity

Physical activity variables demonstrated right-skewed distributions and were therefore log- transformed prior to modelling (log [MET + 1]). Median LTPA_V_ was similar in men and women (1,770 MET-min/week [IQR 813–3,546] and 1,733 MET-min/week [IQR 810– 3,375]), respectively. Men reported higher volumes of vigorous activity than women, while walking contributed the largest proportion of LTPA_V_ in both sexes (Figure 2, Panel B).

Profiles and distributions of LTPA_V_ variables before and after log transformation are shown in Supplementary Information (Table S5, Figure S2).

### LTPA_V_ and odds of fracture

LTPA_V_ demonstrated a non-linear association with odds of fracture (P for non-linearity <0.001), with predicted odds of fracture beginning to increase at approximately 1,800 MET- minutes/week (back transformed from the modelled log scale) (Supplementary Table S6/Supplementary Figure S3). LTPA_V_ was therefore categorised into quintiles for subsequent analyses. Findings were similar in sensitivity analyses using days/week of activity, a variable with lower missing data (Supplementary Figure S4). There was evidence of interaction between LTPA_V_ and both sex and age based on likelihood ratio tests (p <0.001), and analyses were therefore stratified accordingly. Supplementary Table S7 (age and sex interactions)

In women, fracture odds remained close to the null across low-to-moderate quintiles of LTPA_V_ (Q2-3) and increased only in the higher activity quintiles (Q4–Q5) (Figure 3, Supplementary Table S8). The strongest association was observed in women aged 40–49 years (Q5 OR 1.42, 95% CI 1.28–1.58 fully adjusted model), whereas little evidence of association was observed among women aged 60–65 years. In men, associations were stronger and demonstrated a graded dose–response pattern, particularly among those aged 40–59 years (Figure 2, Panel B). The strongest association was observed in men aged 40–49 years (Q5 OR 1.81, 95% CI 1.64–1.99 fully adjusted model).

**Figure 3:**
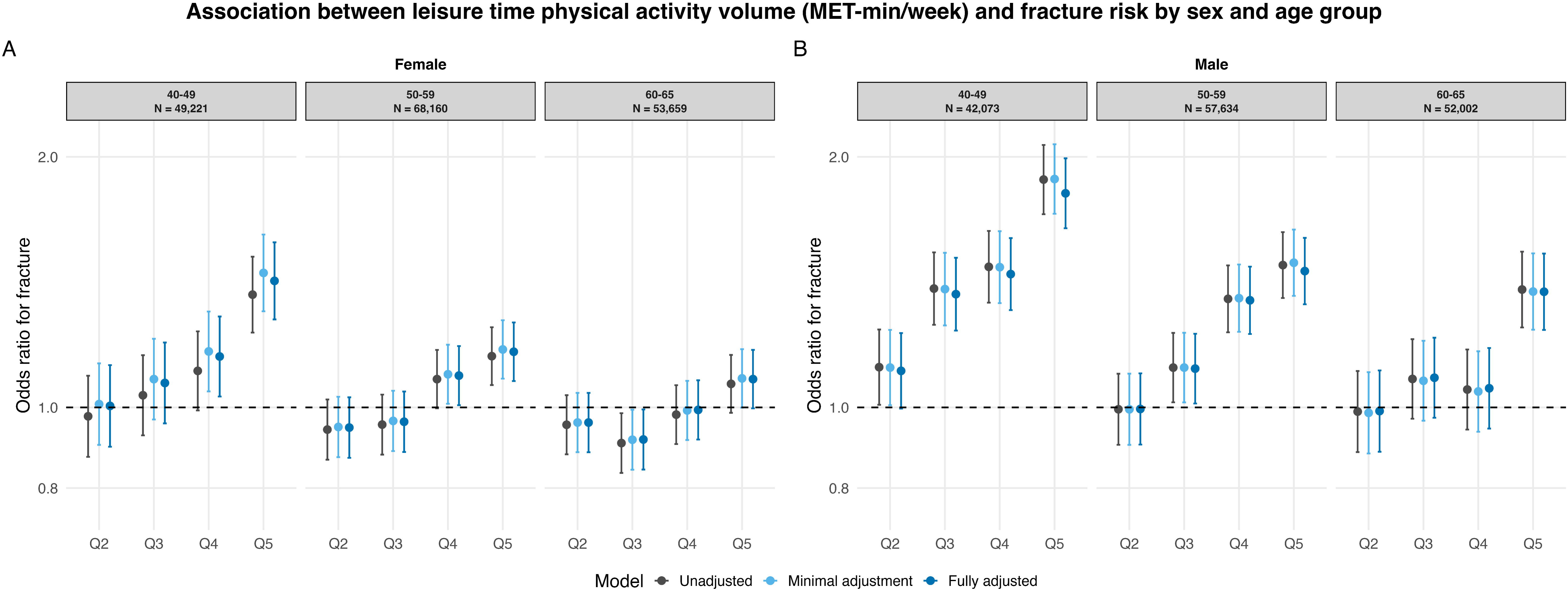
**Associations between leisure-time physical activity volume (LTPA_V_) quintiles and self-reported fracture by sex and age group**. Odds ratios (95% confidence intervals) are shown for activity quintiles Q2–Q5 relative to the lowest activity quintile (Q1; reference category). Estimates are presented for unadjusted, minimally adjusted and fully adjusted models.

Although odds of fracture increased at the highest quintile of LTPA_V_, the absolute differences were small. In women, fracture prevalence was 11.0 per 100 in quintile 5 of LTPA compared with 9.3 per 100 in quintile 1, and in men 11.2 per 100 compared with 7.4 per 100. Estimates were similar across adjustment models. Findings were unchanged when alternative measures of adiposity, including BMI and waist circumference, were used in place of height and weight (Supplementary Table S4, Figure S5).

### Activity intensity and odds of fracture

In mutually adjusted models, associations differed by activity intensity and sex (Figure 4). Among women, there was little evidence of increased fracture odds across most walking, moderate, or vigorous activity quintiles. Modest positive associations were observed only among women aged 40-49 years in the highest quintile of vigorous activity (OR 1.27, 95% CI 1.13–1.42) and moderate activity (OR 1.23, 95% CI 1.09–1.39). In men, vigorous activity demonstrated the strongest and most consistent association with fracture odds increasing progressively across vigorous activity quintiles. The association was strongest with men aged 40-49 years, where those in the highest quintile had 78% higher odds of fracture than those in the lowest quintile (OR 1.78 (1.61-1.98)). Positive associations were also observed among men aged 50–59 years (OR 1.45, 95% CI 1.34–1.57) and 60–65 years (OR 1.25, 95% CI 1.14–1.38). In contrast, associations for walking activity were generally weak and inconsistent. Full results are presented in Supplementary Table S9.

**Figure 4:**
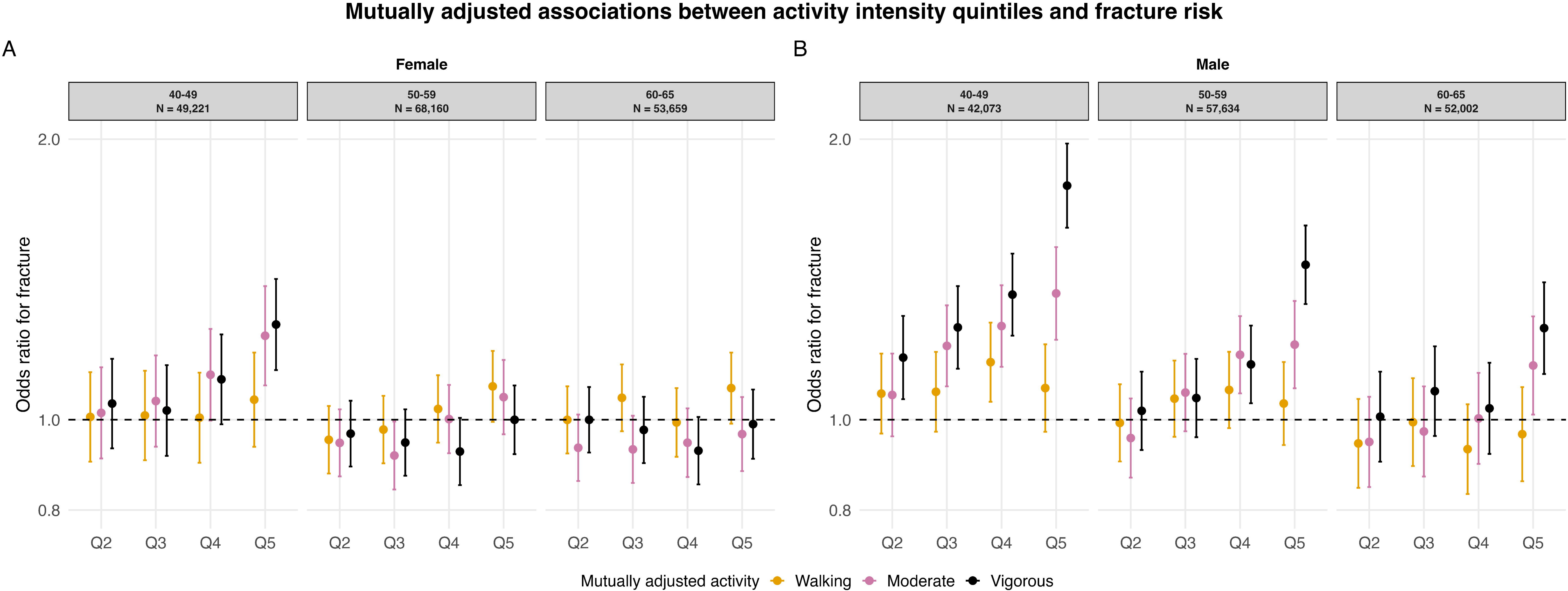
Mutually adjusted associations between activity intensity and odds of fracture by sex and age group. Odds ratios (95% confidence intervals) for fracture according to quintiles of walking, moderate and vigorous intensity physical activity volume (MET minutes/week) in mutually adjusted models, stratified by sex and age group. Models included walking, moderate and vigorous activity simultaneously. Q1 represents the lowest activity quintile and serves as the reference.

Sensitivity analyses examining activity intensity distribution while holding total activity volume constant were consistent with the main findings. There was little evidence of association in women, whereas in men a greater proportion of vigorous activity was associated with higher fracture odds and a greater proportion of walking activity with lower fracture odds at equivalent total activity volumes (Supplementary Figure S6; Supplementary Table S10).

### WHO guideline-defined activity and odds of fracture

Women whose physical activity levels were between the lower and upper targets recommended by the WHO showed a similar fracture odds to women whose activity levels failed to reach the lower target (Figure 5). Amongst women whose physical activity levels exceeded the upper target, a modest increase in fracture odds was observed in those aged 40–49 years (OR 1.19, 95% CI 1.09–1.31). Men whose physical activity levels were between the lower and upper targets recommended by the WHO also showed a similar fracture odds to those whose activity levels failed to reach the lower target. Men whose physical activity levels exceeded the upper target showed an increase in odds of fracture across age groups, which was most marked in those age 40-49 years (Figure ). Full effect estimates are in Supplementary Table S11).

**Figure 5:**
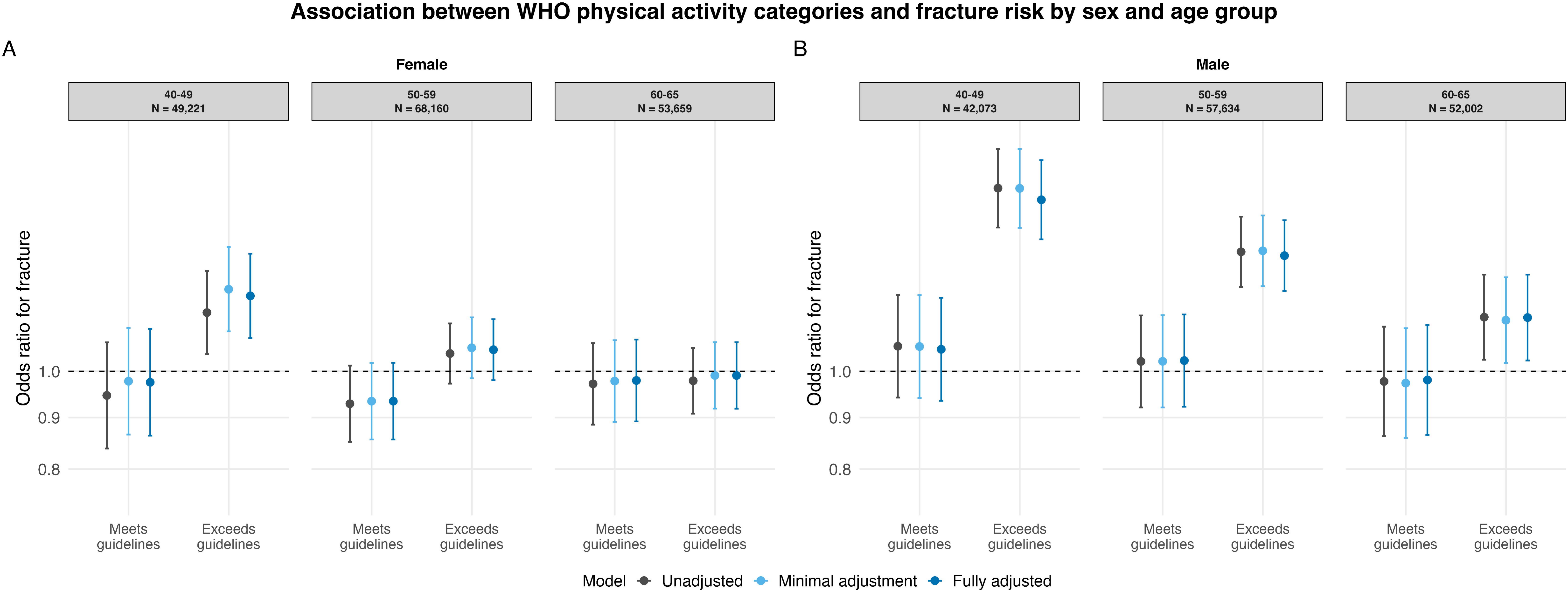
Association between physical activity guideline categories and odds of fracture by age group and sex. Odds ratios and 95% confidence intervals for fracture according to whether World Health Organization (WHO) guidelines for physical activity have been met or exceeded compared to a baseline of not met. Results are shown for unadjusted, minimally adjusted and fully adjusted models. Stratified by age group and sex.

## DISCUSSION

This study examined associations between volumes of LTPA, activity intensity and odds of fracture during mid-life. Importantly physical activity volumes between the lower and higher targets for WHO recommendations were not associated with increased odds of fracture in either sex, supporting continued promotion of physical activity during mid-life. LTPA_V_ beyond the higher target was associated with increased fracture odds, although this relationship was non-linear and concentrated among those undertaking the highest activity volumes, associations being strongest in men, younger adults and for vigorous activity.

A notable finding of this study was that increased fracture odds in women were evident only at the higher quintiles of LTPA_V_. Only two previous studies have examined very high levels of physical activity, reporting either a reduced risk of hip fracture^30^ or no association of extent of physical activity with overall odds of fracture.^31^ Differences between these findings and the current study may reflect variation in fracture outcomes, populations and exposure assessment. Hip fracture risk may be more strongly influenced by the long-term skeletal benefits of physical activity, whereas our outcome included fractures across anatomical sites, many of which may result from increased exposure to injury at higher activity volumes. In our study men demonstrated a more graded relationship between LTPA_V_ and odds of fracture, with fracture odds increasing progressively across activity quintiles and higher risk being observed at lower activity levels than observed in women.

Mutually adjusted analyses indicated that vigorous activity demonstrated the strongest and most consistent independent association with odds of fracture, particularly in men. Although overall LTPAV was broadly similar between the sexes, men reported higher volumes of vigorous activity. Differences in both the volume and context of vigorous activity may therefore contribute to the observed sex differences in fracture odds.

This interpretation is supported by evidence suggesting that women are more likely to participate in structured exercise (e.g. gym classes), and social physical activities, whereas men are more likely to engage in competitive and team based sports.^32,33^ Consequently, similar volumes of vigorous activity in men and women may reflect different patterns of injury exposure.

Competitive and team-based sports typically involve greater opportunities for contact, collisions and falls than many forms of structured exercise,^34,35^ potentially contributing to the higher odds of fracture observed in men. UKB does not record injury mechanism, however, sports participation has been reported to be the third most common cause of fracture in adults, occurs more frequently in men, and predominantly results in fracture of the wrist and hand.^35,36^ This is broadly consistent with the fracture distribution in our cohort which showed that wrist fractures were the most common site-specific fracture type. The relative scarcity of common fragility fractures such as hip suggests that many fractures observed in this cohort of individuals between age 40-65 may be attributable to traumatic injury mechanisms rather than skeletal vulnerability. This interpretation is consistent with previous reports of increased fracture risk among adolescent and young adult males participating in sport,^4^ suggesting that similar mechanisms may persist into mid-life.

Our findings should not be interpreted as suggesting that lower volume or lower-intensity LTPA is universally preferable, particularly in men. Higher volumes and intensities of LTPA, including those exceeding current WHO recommendations, are associated with additional health benefits,^37^ however our findings suggest that these may in part be offset by increased odds of fracture.

### Clinical and Policy Implications

From a public health perspective our study provides reassurance that WHO recommended volumes of LTPA are not associated with increased odds of fracture in women across mid-life and given the substantial wider health benefits of physical activity, it is important that concerns regarding odds of fracture do not unnecessarily discourage physical activity participation, particularly around the menopause. Given the increasing emphasis on exercise as a component of menopause management, these findings could contribute to future updates on safe volumes and intensities of activity in NICE menopause guidance^19^

For health practitioners working with highly active individuals, greater emphasis should be placed on injury prevention and safe participation. Individuals seeking the benefits of vigorous activity should consider how it is accumulated, as different sports and activities may carry different injury risks.

### Strengths and Limitations

A key strength of this study is its large sample size, which enabled stratified analyses by age and sex and detailed examination of activity volume and intensity across mid-life. Self-reported fracture ascertainment enabled inclusion of fractures that may not result in hospital admission, an important consideration in a middle-aged population. However, the cross-sectional design presents an important limitation: physical activity was reported for the preceding seven days, whereas fractures were reported for the preceding five years.

Consequently, we cannot establish whether reported activity preceded fracture, or whether previous fracture influenced subsequent activity levels (reverse causality). Current activity may also not reflect habitual activity during the period in which fractures occurred. These findings should therefore be interpreted as associations rather than evidence that physical activity increases odds of fracture.

Both physical activity and fracture were self-reported, introducing potential recall and reporting bias. Physical activity may be over- or underestimated, potentially leading to misclassification of activity categories and either attenuation or distortion of observed associations. Fractures may be forgotten or unrecognised, particularly clinically silent vertebral fractures; if under-reporting differs by activity level, the direction of bias is uncertain. Information on activity type, fracture mechanism and relevant prior health or functional status was limited, and residual confounding by factors such as comorbidity, medication use and physical function cannot be excluded. Finally, UK Biobank participants are not fully representative of the UK population, potentially limiting generalisability.

## Conclusion

Higher LTPA_V_ in mid-life was associated with higher odds of reporting a fracture, but this was largely confined to individuals undertaking the highest activity volumes. The association was strongest in men and younger middle-aged adults and appeared to be driven, at least in part, by vigorous activity. The findings suggest that activity context, in addition to activity volume, may contribute to odds of fracture, particularly among men. Importantly, physical activity levels consistent with current WHO recommendations were not associated with increased odds of fracture in either sex, supporting continued promotion of physical activity during mid-life while informing future guidance on activity volume and intensity.

## List of abbreviations

CI: Confidence interval
IQR: Inter quartile range
LTPA: Leisure-time physical activity
LTPA_V_: Leisure-time physical activity volume
MET: Metabolic equivalent of task
OR: Odds ratio
Q: Quintile
SD: Standard deviation
UKB: UK Biobank
WHO: World Health Organization

## Declarations

### Ethics approval:/Consent for publication

UK Biobank received ethical approval from the North West Multi-centre Research Ethics Committee (MREC) (reference 11/NW/0382). All participants provided written informed consent. This research was conducted using the UK Biobank resource under application number 81499.

### Equity, diversity, inclusion

This study used data for UKB, a large prospective cohort study with known limitations in representativeness being more White, educated and less deprived than the UK population.^38^ The study team was gender diverse with a balance of clinicians and academics.

### Data Availability Statement

The data analysed in this study are available from the UK Biobank website with approved access. Full code and outputs related to this analysis is available at GitHub (https://github.com/RollsCf/UKB_PA_Fracture.git)

### Patient and Public Involvement

Patients and members of the public were not involved in this study, although UKB is a cohort established through the voluntary participation of members of the public.

### Authors’ contributions

CR analysed and interpreted the data and was a major contributor in writing the manuscript. JT contributed to analysis of findings and manuscript preparation and proofing, HD contributed to data analyses and manuscript proofing, MM and EC were instrumental in study conception and manuscript proofing. BF contributed to study design, code analyses, data interpretation and contributed to manuscript preparation and proofing.

## Acknowledgements

Not applicable

## References

1. Bonafede M, Espindle D, Bower AG. The direct and indirect costs of long bone fractures in a working age US population. J Med Econ 2013; 16: 169–178.

2. Yilmaz ET, Kamaci S, Bingol I, et al. Fracture analysis of working-age adults in Turkey: a 7-year national registry study. BMC Musculoskeletal Disorders 2025; 26: 359.

3. Levy JF, Reider L, Scharfstein DO, et al. The 1-Year Economic Impact of Work Productivity Loss Following Severe Lower Extremity Trauma. J Bone Joint Surg Am 2022; 104: 586–593.

4. Clark EM, Ness AR, Tobias JH. Vigorous Physical Activity Increases Fracture Risk in Children Irrespective of Bone Mass: A Prospective Study of the Independent Risk Factors for Fractures in Healthy Children. J Bone Miner Res 2008; 23: 1012–1022.

5. Rolls C, Dawes H, Munafo M, et al. Epidemiology of mid-life fracture: Self-Reported and Hospital Episode Data from the UK Biobank. J Bone Miner Res 2026; zjag046.

6. Shieh A, Karlamangla AS, Karvonen-Guttierez C, et al. Menopause-related changes in body composition are associated with subsequent bone mineral density and fractures: Study of Women’s Health Across the Nation. J Bone Miner Res 2023; 38: 395–402.

7. Greendale GA, Jackson NJ, Shieh A, et al. Leisure time physical activity and bone mineral density preservation during the menopause transition and postmenopause: a longitudinal cohort analysis from the Study of Women’s Health Across the Nation (SWAN). The Lancet Regional Health - Americas 2023; 21: 100481.

8. Cauley JA, Danielson ME, Greendale GA, et al. Bone resorption and fracture across the menopausal transition: the Study of Women’s Health Across the Nation. Menopause 2012; 19: 1200–1207.

9. Greendale GA, Han W, Huang M, et al. Longitudinal Assessment of Physical Activity and Cognitive Outcomes Among Women at Midlife. JAMA Network Open 2021; 4: e213227.

10. Nguyen B, Clare P, Mielke GI, et al. Physical activity across midlife and health-related quality of life in Australian women: A target trial emulation using a longitudinal cohort. PLoS Med 2024; 21: e1004384.

11. Luo M, Clare PJ, Tarp J, et al. Dose-response interplay between light and moderate-to-vigorous physical activity on all-cause mortality risk: a causal inference analysis. Br J Sports Med. Epub ahead of print 23 March 2026. DOI: 10.1136/bjsports-2025-110782.

12. Chudasama YV, Khunti KK, Zaccardi F, et al. Physical activity, multimorbidity, and life expectancy: a UK Biobank longitudinal study. BMC Medicine 2019; 17: 108.

13. Ramakrishnan R, Doherty A, Smith-Byrne K, et al. Accelerometer measured physical activity and the incidence of cardiovascular disease: Evidence from the UK Biobank cohort study. PLOS Medicine 2021; 18: e1003487.

14. Shreves AH, Small SR, Walmsley R, et al. Amount and intensity of daily total physical activity, step count and risk of incident cancer in the UK Biobank. Br J Sports Med 2025; 59: 839–847.

15. Stattin K, Höijer J, Hållmarker U, et al. Fracture risk across a wide range of physical activity levels, from sedentary individuals to elite athletes. Bone 2021; 153: 116128.

16. Reventlow SD. Perceived risk of osteoporosis: restricted physical activities? Qualitative interview study with women in their sixties. Scand J Prim Health Care 2007; 25: 160– 165.

17. Simmonds BAJ, Hannam KJ, Fox KR, et al. An exploration of barriers and facilitators to older adults’ participation in higher impact physical activity and bone health: a qualitative study. Osteoporos Int 2016; 27: 979–987.

18. Brooke-Wavell K, Skelton DA, Barker KL, et al. Strong, steady and straight: UK consensus statement on physical activity and exercise for osteoporosis. Br J Sports Med 2022; 56: 837–846.

19. National Institue for Health and Care Excellence. National Institute for Health and Care Excellence (2024) Menopause: identification and management, https://www.nice.org.uk/guidance/ng23 (2024).

20. Rolls C, Bastiani O, Antuna M, et al. What is the association between physical activity and fracture risk in middle-aged adults (aged 30-60): a systematic review and synthesis without meta-analysis (SWIM). Osteoporos Int 2026; 37: 57–68.

21. Karlsson M. Does exercise reduce the burden of fractures? A review. Acta Orthop Scand 2002; 73: 691–705.

22. Montgomery G, Yusuf M, Cooper R, et al. Are associations between physical activity and bone mineral density in adults sex- and age-dependent? An analysis of the UK Biobank study. Journal of Bone and Mineral Research 2024; 39: 399–407.

23. Sudlow C, Gallacher J, Allen N, et al. UK Biobank: An Open Access Resource for Identifying the Causes of a Wide Range of Complex Diseases of Middle and Old Age. PLOS Medicine 2015; 12: e1001779.

24. IPAQ Research Committee. Guidelines for Data Processing and Analysis of the International Physical Activity Questionnaire (IPAQ)-Short and Long Forms. http://www.ipaq.ki.se/scoring.pdf, https://cir.nii.ac.jp/crid/1573950400545533440 (2005, accessed 19 March 2024).

25. Townsend P, Phillimore P, Beattie A. Health and Deprivation: Inequality and the North. London: Routledge, 1988. Epub ahead of print 1988. DOI: 10.4324/9781003368885.

26. UK Biobank. UK Biobank. Anthropometry (Assessment Centre) Standard Operating Procedure., https://biobank.ctsu.ox.ac.uk/crystal/crystal/docs/Anthropometry.pdf (2007).

27. Vandenbroucke JP, von Elm E, Altman DG, et al. Strengthening the Reporting of Observational Studies in Epidemiology (STROBE): explanation and elaboration. Epidemiology 2007; 18: 805–835.

28. Mansournia MA, Collins GS, Nielsen RO, et al. CHecklist for statistical Assessment of Medical Papers: the CHAMP statement. Epub ahead of print 1 September 2021. DOI: 10.1136/bjsports-2020-103651.

29. Rolls C, Tobias J, Munafo M, et al. OSF Registration: Exploring the association and temporal relationships between midlife physical activity and fracture risk, https://osf.io/4g87k (2026, accessed 28 April 2026).

30. Feskanich D, Willett W, Colditz G, et al. Walking and leisure-time activity and risk of hip fracture in postmenopausal women. JAMA: Journal of the American Medical Association 2002; 288: 2300–2306.

31. Jordan S, Lim L, Berecki-Gisolf J, et al. Body mass index, physical activity, and fracture among young adults: longitudinal results from the Thai cohort study. J Epidemiol 2013; 23: 435–42.

32. Owen KB, Clare PJ, Eime R, et al. Gender gap in physical activity and sport participation across the lifespan in Australia between 2016 and 2023. BMC Public Health 2025; 25: 2889.

33. Owen KB, Corbett L, Ding D, et al. Gender differences in physical activity and sport participation in adults across 28 European countries between 2005 and 2022. Ann Epidemiol 2025; 101: 52–57.

34. Crossman S, Drummond M, Elliott S, et al. Facilitators and constraints to adult sports participation: A systematic review. Psychology of Sport and Exercise 2024; 72: 102609.

35. Court-Brown CM, Wood AM, Aitken S. The epidemiology of acute sports-related fractures in adults. Injury 2008; 39: 1365–1372.

36. Meixner C, Loder RT. The Demographics of Fractures and Dislocations Across the Entire United States due to Common Sports and Recreational Activities. Sports Health 2019; 12: 159–169.

37. Garcia L, Pearce M, Abbas A, et al. Non-occupational physical activity and risk of cardiovascular disease, cancer and mortality outcomes: a dose–response meta-analysis of large prospective studies. Br J Sports Med 2023; 57: 979–989.

38. Fry A, Littlejohns TJ, Sudlow C, et al. Comparison of Sociodemographic and Health-Related Characteristics of UK Biobank Participants With Those of the General Population. Am J Epidemiol 2017; 186: 1026–1034.

