## supplementary information for "HOW SAFE IS EXERCISE? CROSS-SECTIONAL ASSOCIATIONS OF CURRENT PHYSICAL ACTIVITY WITH SELF-REPORTED FRACTURE HISTORY IN MID-LIFE"

#### Table of Contents

**Supplementary Methods**

S1: Changes to the analysis plan

S2: Classification of meeting or exceeding WHO recommendations

**Supplementary Tables**

Table S1: Response rate and percentage missingness

Table S2: Comparison of those with and without activity data and standardised mean differences.

Table S3 Activity volume by age, sex and intensity

Table S4: Assessment of linearity for continuous physical activity measures (likelihood ratio test comparing linear and spline models)

Table S5: Assessment of sex and age-group Likelihood Ratio Test (LRT) interaction for spline physical activity models

Table S6: Odds ratios for total physical activity quintiles and self-reported fracture, stratified by sex and age group

Table S7: Mutually adjusted activity (Odds Ratio and 95% CI)

Table S8: Association between a 10% higher proportion of activity type within a fixed total activity volume and fracture risk (OR, 95% CI)

Table S9: Adjusted odds ratios for WHO physical activity categories and self-reported fracture, stratified by sex

**Supplementary Figures**

Supplementary Figure S2: Distribution of MET variables before and after log transformation (Log+1)

Supplementary Figure S3: Association between Total activity (MET log+1) and Fracture

Supplementary Figure S4: Sensitivity analysis of the association between total physical activity frequency (days/week) and self-reported fracture by sex and age group.

Supplementary Figure S5. Association between total physical activity and self-reported fracture using alternative adiposity adjustment specifications.

Supplementary Figure S6. Association between activity composition and fracture risk within total physical activity quintiles

#### Supplementary Methods

##### Supplementary Methods S1

Physical Activity Questionnaire (IPAQ). We will examine physical activity using the following pre-specified measures: Categorical IPAQ physical activity groups (low, moderate, high), defined according to IPAQ scoring guidelines.

| | |
|---|---|
| Plan | Physical Activity Questionnaire (IPAQ). We will examine physical activity using the following pre-specified measures: Categorical IPAQ physical activity groups (low, moderate, high), defined according to IPAQ scoring guidelines.<br><br>In addition, we will conduct sensitivity analyses examining the association between outcomes and meeting or exceeding the World Health Organization physical activity recommendations for MVPA and walking. |
| Change | We used part two instead of part 1, i.e. WHO guidelines instead of IPAQ in order to understand public health impact |
| Impact | Moderate: Improved ability to make public health recommendations. |

| | |
|---|---|
| Plan | Fracture ascertainment: *Self-reported fracture in mid-life*. The primary outcomes will be self-reported hip fracture due to the high mortality and morbidity associated with this fracture and self-reported wrist fracture due to both the cost economic impact of this fracture in working aged adults and their role as an early indicator of skeletal fragility. Both will be taken from assessment centre 0. A sensitivity analysis will be conducted on all fracture. |
| Change | We used total fracture in the cross-sectional analysis. This was to increase power to detect meaningful change. We elected to use site specific fracture in the parallel survival analyses specified in the analysis plan as sample size is larger with longer follow-up |
| Impact | Low |

**Supplementary Methods S2. WHO Physical Activity Categories**

To contextualise findings relative to current public health recommendations, participants were additionally categorised according to whether they did not meet, met, or exceeded the World Health Organization (WHO) physical activity guidelines for adults.

Weekly moderate-equivalent physical activity was calculated as the sum of weekly walking minutes, moderate-intensity activity minutes, and twice the weekly vigorous-intensity activity minutes (walking + moderate + [2 × vigorous]). This approach reflects the WHO recommendation that 1 minute of vigorous-intensity activity provides approximately equivalent health benefit to 2 minutes of moderate-intensity activity.

Participants were classified as:

- **Does not meet guidelines:** <150 minutes/week of moderate-equivalent activity
- **Meets guidelines:** 150–300 minutes/week of moderate-equivalent activity
- **Exceeds guidelines:** >300 minutes/week of moderate-equivalent activity

These categories correspond to the lower and upper thresholds of the WHO recommendation for weekly moderate-intensity physical activity, allowing findings to be interpreted in relation to established public health guidance.

### Supplementary Tables

*Supplementary Table S1: Response rate and percentage missing data by study variable*

| Variable | Answered (N) | Answered (%) | Missing (N) | Missing (%) |
|---|---|---|---|---|
| Ethnicity | 425737 | 99 | 2338 | 0.55 |
| Fracture | 424727 | 99 | 3348 | 0.78 |
| Number of days of moderate activity | 406360 | 95 | 21715 | 5.07 |
| Number of days walking | 419567 | 98 | 8508 | 1.99 |
| Number of days vigorous activity | 406654 | 95 | 21421 | 5.00 |
| Moderate activity (MET minutes/week) | 368265 | 86 | 59810 | 13.97 |
| Vigorous activity (MET minutes/week) | 383931 | 90 | 44144 | 10.31 |
| Walking activity (MET minutes/week) | 372667 | 87 | 55408 | 12.94 |
| LTPA volume (MET minutes/week) | 332403 | 78 | 95672 | 22.35 |
| Education | 415537 | 97 | 12538 | 2.93 |
| Townsend Deprivation Index | 427501 | 99 | 574 | 0.13 |
| Weight | 425715 | 99 | 2360 | 0.55 |
| Height | 425909 | 99 | 2166 | 0.51 |
| BMI | 425441 | 99 | 2634 | 0.62 |

LTPA = Leisure Time Physical Activity, MET = Metabolic Equivalent Task unit, BMI = Body Mass Index

*Supplementary Table S2: Comparison of those with and without activity data and standardised mean differences.*

| Variable | **Overall** N = 428,075[1] | **Has activity data** N = 332,403[1] | **Missing activity data** N = 95,672[1] | **SMD**[2] |
|---|---|---|---|---|
| Sex, n (%) | | | | 0.20 |
| Female | 235,713 (55%) | 175,795 (53%) | 59,918 (63%) | |
| Male | 192,362 (45%) | 156,608 (47%) | 35,754 (37%) | |
| Age (years), mean (SD) | 54.6 (7.2) | 54.4 (7.3) | 55.4 (7.1) | 0.14 |
| Ethnicity, n (%) | | | | 0.11 |
| Asian | 8,875 (2.1%) | 5,970 (1.8%) | 2,905 (3.1%) | |
| Black or other | 13,060 (3.1%) | 9,205 (2.8%) | 3,855 (4.1%) | |
| Mixed | 2,747 (0.6%) | 2,119 (0.6%) | 628 (0.7%) | |
| White | 401,055 (94%) | 314,170 (95%) | 86,885 (92%) | |
| Townsend deprivation index, mean (SD) | -1.3 (3.1) | -1.4 (3.0) | -0.8 (3.3) | 0.18 |
| Education level, n (%) | | | | 0.05 |
| School education or less | 350,239 (84%) | 274,673 (84%) | 75,566 (85%) | |
| College or vocational training | 39,402 (9.5%) | 31,126 (9.5%) | 8,276 (9.3%) | |
| University degree or above | 25,896 (6.2%) | 21,135 (6.5%) | 4,761 (5.4%) | |
| Weight (kg), mean (SD) | 78.2 (16.2) | 78.3 (16.1) | 77.8 (16.6) | 0.03 |
| Height (cm), mean (SD) | 168.7 (9.3) | 169.2 (9.3) | 166.7 (9.1) | 0.28 |
| BMI (kg/m²), mean (SD) | 27.4 (4.9) | 27.3 (4.7) | 28.0 (5.3) | 0.14 |
| Any fracture, n (%) | | | | 0.02 |
| No | 384,525 (91%) | 299,896 (91%) | 84,629 (90%) | |
| Yes | 40,202 (9.5%) | 30,926 (9.3%) | 9,276 (9.9%) | |

[1]n (%); Mean (SD)

[2]SMD = standardised mean difference. Values <0.1 indicate negligible differences, 0.1–0.2 small differences, and >0.2 meaningful imbalance.

*Table S3: Distribution of fracture sites by age group and sex*

| sex | agegp_A0 | Ankle | Arm | Hip | Leg | Other | Spine | Wrist |
|---|---|---|---|---|---|---|---|---|
| Female | 40-49 | 505 (13.1%) | 328 (8.5%) | 24 (0.6%) | 179 (4.6%) | 2167 (56.2%) | 77 (2%) | 578 (15%) |
| Female | 50-59 | 1001 (14.3%) | 731 (10.5%) | 86 (1.2%) | 361 (5.2%) | 3325 (47.6%) | 152 (2.2%) | 1324 (19%) |
| Female | 60-65 | 1128 (15.4%) | 821 (11.2%) | 138 (1.9%) | 419 (5.7%) | 2853 (38.8%) | 143 (1.9%) | 1845 (25.1%) |
| Male | 40-49 | 552 (10.1%) | 456 (8.3%) | 49 (0.9%) | 349 (6.4%) | 3356 (61.1%) | 88 (1.6%) | 639 (11.6%) |
| Male | 50-59 | 580 (10.4%) | 484 (8.6%) | 88 (1.6%) | 394 (7%) | 3244 (57.9%) | 103 (1.8%) | 707 (12.6%) |
| Male | 60-65 | 458 (11.7%) | 367 (9.4%) | 95 (2.4%) | 301 (7.7%) | 2014 (51.5%) | 96 (2.5%) | 583 (14.9%) |

*Table S4: Total fractures per 100 participants by leisure time physical activity volume quintile*

| sex | Activity quintile | N | Total fracture, n (%) |
|---|---|---|---|
| Female | Q1 | 33904 | 3159 (9.3%) |
| Female | Q2 | 35526 | 3226 (9.1%) |
| Female | Q3 | 35098 | 3212 (9.2%) |
| Female | Q4 | 34689 | 3464 (10.0%) |
| Female | Q5 | 31823 | 3501 (11.0%) |
| Male | Q1 | 30646 | 2275 (7.4%) |
| Male | Q2 | 29024 | 2217 (7.6%) |
| Male | Q3 | 29452 | 2564 (8.7%) |
| Male | Q4 | 29861 | 2820 (9.4%) |
| Male | Q5 | 32726 | 3650 (11.2%) |

*Supplementary Table S5 Activity volume by Age, sex and intensity*

| Age group | Sex | PA quintile | N | LTPAv MET-min/week | LTPA min/week | Walking min/week | Moderate min/week | Vigorous min/week |
|---|---|---|---|---|---|---|---|---|
| 40-49 | Female | Q1 | 9977 | 339 (179–495) | 90 (50–135) | 60 (30–100) | 10 (0–30) | 0 (0–0) |
| | | Q2 | 10259 | 982 (815–1160) | 240 (205–290) | 150 (90–210) | 60 (30–90) | 16 (0–45) |
| | | Q3 | 10427 | 1773 (1542–2026) | 420 (360–480) | 210 (120–360) | 100 (60–180) | 60 (6–120) |
| | | Q4 | 10039 | 3039 (2634–3511) | 710 (600–850) | 350 (210–560) | 180 (120–300) | 100 (30–180) |
| | | Q5 | 8519 | 5918 (4893–7758) | 1470 (1200–1860) | 840 (420–1260) | 480 (300–840) | 180 (60–300) |
| | Male | Q1 | 8113 | 330 (148–490) | 85 (40–125) | 50 (20–100) | 10 (0–30) | 0 (0–10) |
| | | Q2 | 7903 | 990 (815–1170) | 235 (195–280) | 140 (70–210) | 50 (20–90) | 30 (0–60) |
| | | Q3 | 8350 | 1782 (1546–2031) | 405 (330–470) | 180 (100–315) | 90 (45–150) | 80 (20–120) |
| | | Q4 | 8365 | 3060 (2648–3546) | 675 (560–810) | 280 (150–420) | 180 (100–300) | 135 (60–210) |
| | | Q5 | 9342 | 6558 (5160–9198) | 1560 (1200–2160) | 840 (420–1260) | 600 (300–900) | 240 (120–420) |
| 50-59 | Female | Q1 | 14691 | 344 (179–495) | 95 (50–135) | 60 (30–100) | 10 (0–30) | 0 (0–0) |
| | | Q2 | 14654 | 975 (806–1158) | 240 (210–300) | 150 (90–210) | 60 (30–100) | 10 (0–40) |
| | | Q3 | 13763 | 1760 (1536–2013) | 430 (370–500) | 210 (140–360) | 120 (60–180) | 40 (0–90) |
| | | Q4 | 13059 | 3057 (2628–3546) | 720 (620–870) | 360 (210–560) | 210 (120–360) | 80 (10–160) |
| | | Q5 | 11993 | 5964 (4932–7776) | 1500 (1230–1890) | 840 (420–1260) | 600 (315–900) | 135 (40–300) |
| | Male | Q1 | 12646 | 330 (148–488) | 85 (40–125) | 52 (20–100) | 10 (0–30) | 0 (0–2) |
| | | Q2 | 11195 | 982 (810–1158) | 240 (205–290) | 150 (90–210) | 60 (20–90) | 15 (0–45) |
| | | Q3 | 11025 | 1770 (1544–2028) | 420 (355–480) | 210 (120–360) | 100 (60–180) | 60 (5–120) |
| | | Q4 | 10917 | 3051 (2634–3546) | 705 (590–840) | 315 (175–455) | 210 (120–300) | 120 (30–180) |
| | | Q5 | 11851 | 6492 (5115–8958) | 1560 (1230–2110) | 840 (420–1260) | 630 (360–900) | 210 (75–390) |
| 60-65 | Female | Q1 | 9236 | 368 (198–504) | 100 (60–140) | 60 (30–100) | 12 (0–40) | 0 (0–0) |
| | | Q2 | 10613 | 978 (813–1160) | 250 (210–300) | 140 (90–210) | 75 (40–120) | 0 (0–30) |
| | | Q3 | 10908 | 1773 (1533–2026) | 435 (380–510) | 210 (135–350) | 150 (90–225) | 30 (0–80) |
| | | Q4 | 11591 | 3066 (2651–3546) | 740 (630–870) | 315 (200–420) | 300 (180–420) | 60 (0–135) |
| | | Q5 | 11311 | 5958 (4932–7758) | 1470 (1200–1860) | 630 (420–1080) | 720 (420–900) | 120 (30–270) |
| | Male | Q1 | 9887 | 338 (165–495) | 90 (45–135) | 60 (30–100) | 10 (0–30) | 0 (0–0) |
| | | Q2 | 9926 | 975 (810–1158) | 250 (210–300) | 150 (100–210) | 60 (30–105) | 5 (0–30) |
| | | Q3 | 10077 | 1764 (1533–2013) | 430 (375–500) | 210 (135–360) | 120 (60–210) | 30 (0–90) |
| | | Q4 | 10579 | 3066 (2639–3546) | 724 (620–860) | 315 (180–480) | 240 (140–400) | 80 (10–160) |
| | | Q5 | 11533 | 6318 (5026–8398) | 1500 (1210–1980) | 630 (360–1080) | 720 (420–900) | 180 (60–360) |

*LTPA = Leisure time physical activity, MET = Metabolic Equivalent Task unit*

*Supplementary Table S6: Assessment of linearity for continuous physical activity measures (likelihood ratio test (LRT) comparing linear and spline models)*

| Exposure (MET min/week) | Outcome | LRT p value | Linear Adequate |
|---|---|---|---|
| Moderate Intensity volume | All Fracture | <0.001 | No |
| Vigorous Intensity volume | All Fracture | <0.001 | No |
| Walking volume | All Fracture | <0.001 | No |
| LTPA volume | All Fracture | <0.001 | No |
| Moderate Intensity volume | Hip | 0.012 | No |
| Vigorous Intensity volume | Hip | 0.086 | Yes |
| Walking volume | Hip | 0.021 | No |
| LTPA volume | Hip | 0.021 | No |
| Moderate Intensity | Wrist | <0.001 | No |
| Vigorous Intensity | Wrist | <0.001 | No |
| Walking | Wrist | <0.001 | No |
| LTPA volume | Wrist | <0.001 | No |

Likelihood ratio tests (LRTs) compared models including a linear term with models including restricted cubic spline terms for each continuous physical activity exposure. A significant LRT ($P<0.05$) indicates evidence of a non-linear association and that the spline model provides a better fit than the linear model. LTPA = Leisure time physical activity

*Supplementary Table S7: Assessment of sex and age-group Likelihood Ratio Test (LRT) interaction for spline physical activity model*

| outcome | exposure | interaction | LRT p value | Evidence of interaction |
|---|---|---|---|---|
| SRF | Moderate Intensity volume | Sex | <0.001 | Yes |
| SRF | Moderate Intensity volume | Age group | <0.001 | Yes |
| SRF | Vigorous Intensity volume | Sex | <0.001 | Yes |
| SRF | Vigorous Intensity volume | Age group | <0.001 | Yes |
| SRF | Walking volume | Sex | 0.143 | No |
| SRF | Walking volume | Age group | 0.002 | Yes |
| SRF | LTPA volume | Sex | <0.001 | Yes |
| SRF | LTPA volume | Age group | <0.001 | Yes |

Likelihood ratio tests (LRTs) compared spline models with and without interaction terms between physical activity and sex or age group. A significant LRT ($P<0.05$) indicates evidence that the association between physical activity and fracture risk differs by sex or age group. LTPA = Leisure time physical activity

*Supplementary Table S8: Odds ratios for leisure time physical activity volume quintiles and self-reported fracture, stratified by sex and age group*

| Sex | Age Group | Model | Quintile | Odds ratio | CI lower | CI upper | P value | Global P value |
|---|---|---|---|---|---|---|---|---|
| Female | 40-49 | Unadjusted | 2 | 0.98 | 0.87 | 1.09 | 0.669 | <0.001 |
| | | Unadjusted | 3 | 1.03 | 0.93 | 1.16 | 0.551 | NA |
| | | Unadjusted | 4 | 1.11 | 0.99 | 1.23 | 0.070 | NA |
| | | Unadjusted | 5 | 1.37 | 1.23 | 1.52 | <0.001 | NA |
| | | Minimal adjustment | 2 | 1.01 | 0.90 | 1.13 | 0.874 | <0.001 |
| | | Minimal adjustment | 3 | 1.08 | 0.97 | 1.21 | 0.170 | NA |
| | | Minimal adjustment | 4 | 1.17 | 1.05 | 1.30 | 0.006 | NA |
| | | Minimal adjustment | 5 | 1.45 | 1.30 | 1.61 | <0.001 | NA |
| | | Fully adjusted | 2 | 1.00 | 0.90 | 1.12 | 0.942 | <0.001 |
| | | Fully adjusted | 3 | 1.07 | 0.96 | 1.20 | 0.236 | NA |
| | | Fully adjusted | 4 | 1.15 | 1.03 | 1.29 | 0.013 | NA |
| | | Fully adjusted | 5 | 1.42 | 1.28 | 1.58 | <0.001 | NA |
| Male | | Unadjusted | 2 | 1.12 | 1.01 | 1.24 | 0.036 | <0.001 |
| | | Unadjusted | 3 | 1.39 | 1.26 | 1.54 | <0.001 | NA |
| | | Unadjusted | 4 | 1.48 | 1.34 | 1.63 | <0.001 | NA |
| | | Unadjusted | 5 | 1.88 | 1.71 | 2.07 | <0.001 | NA |
| | | Minimal adjustment | 2 | 1.12 | 1.01 | 1.24 | 0.038 | <0.001 |
| | | Minimal adjustment | 3 | 1.39 | 1.25 | 1.53 | <0.001 | NA |
| | | Minimal adjustment | 4 | 1.47 | 1.33 | 1.63 | <0.001 | NA |
| | | Minimal adjustment | 5 | 1.88 | 1.71 | 2.07 | <0.001 | NA |
| | | Fully adjusted | 2 | 1.11 | 1.00 | 1.23 | 0.057 | <0.001 |
| | | Fully adjusted | 3 | 1.37 | 1.24 | 1.51 | <0.001 | NA |
| | | Fully adjusted | 4 | 1.45 | 1.31 | 1.60 | <0.001 | NA |
| | | Fully adjusted | 5 | 1.81 | 1.64 | 1.99 | <0.001 | NA |
| Female | 50-59 | Unadjusted | 2 | 0.94 | 0.87 | 1.02 | 0.149 | <0.001 |
| | | Unadjusted | 3 | 0.95 | 0.88 | 1.04 | 0.262 | NA |
| | | Unadjusted | 4 | 1.08 | 1.00 | 1.17 | 0.058 | NA |
| | | Unadjusted | 5 | 1.15 | 1.06 | 1.25 | <0.001 | NA |
| | | Minimal adjustment | 2 | 0.95 | 0.87 | 1.03 | 0.205 | <0.001 |
| | | Minimal adjustment | 3 | 0.96 | 0.89 | 1.05 | 0.382 | NA |
| | | Minimal adjustment | 4 | 1.10 | 1.01 | 1.19 | 0.028 | NA |
| | | Minimal adjustment | 5 | 1.17 | 1.08 | 1.27 | <0.001 | NA |

| Sex | Age Group | Model | Quintile | Odds ratio | CI lower | CI upper | P value | Global P value |
|---|---|---|---|---|---|---|---|---|
| | | Fully adjusted | 2 | 0.95 | 0.87 | 1.03 | 0.192 | <0.001 |
| | | Fully adjusted | 3 | 0.96 | 0.88 | 1.04 | 0.353 | NA |
| | | Fully adjusted | 4 | 1.09 | 1.01 | 1.19 | 0.035 | NA |
| | | Fully adjusted | 5 | 1.17 | 1.08 | 1.26 | <0.001 | NA |
| Male | | Unadjusted | 2 | 0.99 | 0.90 | 1.10 | 0.920 | <0.001 |
| | | Unadjusted | 3 | 1.12 | 1.01 | 1.23 | 0.024 | NA |
| | | Unadjusted | 4 | 1.35 | 1.23 | 1.48 | <0.001 | NA |
| | | Unadjusted | 5 | 1.48 | 1.35 | 1.62 | <0.001 | NA |
| | | Minimal adjustment | 2 | 0.99 | 0.90 | 1.10 | 0.919 | <0.001 |
| | | Minimal adjustment | 3 | 1.12 | 1.01 | 1.23 | 0.025 | NA |
| | | Minimal adjustment | 4 | 1.35 | 1.23 | 1.48 | <0.001 | NA |
| | | Minimal adjustment | 5 | 1.49 | 1.36 | 1.64 | <0.001 | NA |
| | | Fully adjusted | 2 | 1.00 | 0.90 | 1.10 | 0.933 | <0.001 |
| | | Fully adjusted | 3 | 1.11 | 1.01 | 1.23 | 0.029 | NA |
| | | Fully adjusted | 4 | 1.34 | 1.23 | 1.48 | <0.001 | NA |
| | | Fully adjusted | 5 | 1.46 | 1.33 | 1.60 | <0.001 | NA |
| Female | 60-65 | Unadjusted | 2 | 0.95 | 0.88 | 1.03 | 0.251 | 0.002 |
| | | Unadjusted | 3 | 0.91 | 0.83 | 0.98 | 0.019 | NA |
| | | Unadjusted | 4 | 0.98 | 0.90 | 1.06 | 0.633 | NA |
| | | Unadjusted | 5 | 1.07 | 0.99 | 1.16 | 0.111 | NA |
| | | Minimal adjustment | 2 | 0.96 | 0.88 | 1.04 | 0.319 | 0.002 |
| | | Minimal adjustment | 3 | 0.91 | 0.84 | 0.99 | 0.035 | NA |
| | | Minimal adjustment | 4 | 0.99 | 0.91 | 1.08 | 0.843 | NA |
| | | Minimal adjustment | 5 | 1.08 | 1.00 | 1.18 | 0.052 | NA |
| | | Fully adjusted | 2 | 0.96 | 0.88 | 1.04 | 0.319 | 0.002 |
| | | Fully adjusted | 3 | 0.92 | 0.84 | 0.99 | 0.037 | NA |
| | | Fully adjusted | 4 | 0.99 | 0.91 | 1.08 | 0.870 | NA |
| | | Fully adjusted | 5 | 1.08 | 1.00 | 1.17 | 0.058 | NA |
| Male | | Unadjusted | 2 | 0.99 | 0.88 | 1.11 | 0.841 | <0.001 |
| | | Unadjusted | 3 | 1.08 | 0.97 | 1.21 | 0.160 | NA |
| | | Unadjusted | 4 | 1.05 | 0.94 | 1.17 | 0.381 | NA |
| | | Unadjusted | 5 | 1.39 | 1.25 | 1.54 | <0.001 | NA |
| | | Minimal adjustment | 2 | 0.99 | 0.88 | 1.10 | 0.795 | <0.001 |
| | | Minimal adjustment | 3 | 1.08 | 0.96 | 1.20 | 0.191 | NA |
| | | Minimal adjustment | 4 | 1.05 | 0.94 | 1.17 | 0.437 | NA |

| Sex | Age Group | Model | Quintile | Odds ratio | CI lower | CI upper | P value | Global P value |
|---|---|---|---|---|---|---|---|---|
| | | Minimal adjustment | 5 | 1.38 | 1.24 | 1.53 | <0.001 | NA |
| | | Fully adjusted | 2 | 0.99 | 0.88 | 1.11 | 0.861 | <0.001 |
| | | Fully adjusted | 3 | 1.09 | 0.97 | 1.21 | 0.146 | NA |
| | | Fully adjusted | 4 | 1.05 | 0.94 | 1.18 | 0.351 | NA |
| | | Fully adjusted | 5 | 1.38 | 1.24 | 1.53 | <0.001 | NA |

CI = Confidence Interval

*Supplementary Table S9: Mutually adjusted associations between activity intensity quintiles and self-reported fracture (odds ratios and 95% confidence intervals)*

| **Sex** | **Age group** | **Exposure** | **Q2** | **Q3** | **Q4** | **Q5** |
|---|---|---|---|---|---|---|
| Female | 40-49 | Vigorous | 1.04 (0.93–1.16) | 1.02 (0.91–1.14) | 1.1 (0.99–1.23) | 1.27 (1.13–1.42) |
| Female | 50-59 | Vigorous | 0.97 (0.89–1.05) | 0.95 (0.87–1.03) | 0.92 (0.85–1) | 1 (0.92–1.09) |
| Female | 60-69 | Vigorous | 1 (0.92–1.08) | 0.98 (0.9–1.06) | 0.93 (0.85–1.01) | 0.99 (0.91–1.08) |
| Male | 40-49 | Vigorous | 1.17 (1.05–1.29) | 1.26 (1.13–1.39) | 1.36 (1.23–1.51) | 1.78 (1.61–1.98) |
| Male | 50-59 | Vigorous | 1.02 (0.93–1.13) | 1.06 (0.96–1.16) | 1.15 (1.04–1.26) | 1.47 (1.33–1.62) |
| Male | 60-69 | Vigorous | 1.01 (0.9–1.13) | 1.07 (0.96–1.2) | 1.03 (0.92–1.15) | 1.25 (1.12–1.4) |
| Female | 40-49 | Moderate | 1.02 (0.91–1.14) | 1.05 (0.94–1.17) | 1.12 (1–1.25) | 1.23 (1.09–1.39) |
| Female | 50-59 | Moderate | 0.94 (0.87–1.03) | 0.92 (0.84–1) | 1 (0.92–1.09) | 1.06 (0.96–1.16) |
| Female | 60-69 | Moderate | 0.93 (0.86–1.01) | 0.93 (0.86–1.01) | 0.94 (0.87–1.03) | 0.97 (0.88–1.06) |
| Male | 40-49 | Moderate | 1.06 (0.96–1.18) | 1.2 (1.09–1.33) | 1.26 (1.14–1.39) | 1.37 (1.22–1.53) |
| Male | 50-59 | Moderate | 0.96 (0.87–1.05) | 1.07 (0.97–1.18) | 1.17 (1.07–1.29) | 1.2 (1.08–1.34) |
| Male | 60-69 | Moderate | 0.95 (0.85–1.06) | 0.97 (0.87–1.09) | 1 (0.9–1.12) | 1.14 (1.01–1.29) |
| Female | 40-49 | Walking | 1.01 (0.9–1.12) | 1.01 (0.9–1.13) | 1 (0.9–1.12) | 1.05 (0.94–1.18) |
| Female | 50-59 | Walking | 0.95 (0.88–1.03) | 0.98 (0.9–1.06) | 1.03 (0.94–1.12) | 1.09 (0.99–1.19) |
| Female | 60-69 | Walking | 1 (0.92–1.09) | 1.06 (0.97–1.15) | 0.99 (0.91–1.08) | 1.08 (0.99–1.18) |
| Male | 40-49 | Walking | 1.07 (0.97–1.18) | 1.07 (0.97–1.18) | 1.15 (1.05–1.27) | 1.08 (0.97–1.2) |
| Male | 50-59 | Walking | 0.99 (0.9–1.09) | 1.05 (0.96–1.16) | 1.08 (0.98–1.18) | 1.04 (0.94–1.15) |
| Male | 60-69 | Walking | 0.94 (0.85–1.05) | 0.99 (0.89–1.11) | 0.93 (0.83–1.04) | 0.96 (0.86–1.08) |

*Supplementary Table S10: Association between a 10% higher proportion of activity type within a fixed total activity volume and fracture risk (OR, 95% CI)*

| model | sex | OR | CI lower | CI upper | P value |
|---:|---:|---:|---:|---:|---:|
| Vigorous Q1 | Female | 1.01 | 0.99 | 1.03 | 0.35 |
| Moderate Q1 | Female | 1.00 | 0.98 | 1.01 | 0.59 |
| Walk Q1 | Female | 1.00 | 0.99 | 1.01 | 0.98 |
| Vigorous Q2 | Female | 0.99 | 0.97 | 1.00 | 0.14 |
| Moderate Q2 | Female | 1.00 | 0.98 | 1.01 | 0.69 |
| Walk Q2 | Female | 1.01 | 1.00 | 1.02 | 0.13 |
| Vigorous Q3 | Female | 0.99 | 0.97 | 1.01 | 0.20 |
| Moderate Q3 | Female | 1.00 | 0.98 | 1.02 | 0.85 |
| Walk Q3 | Female | 1.01 | 1.00 | 1.02 | 0.21 |
| Vigorous Q4 | Female | 1.00 | 0.98 | 1.02 | 0.97 |
| Moderate Q4 | Female | 1.01 | 0.99 | 1.02 | 0.38 |
| Walk Q4 | Female | 0.99 | 0.98 | 1.01 | 0.44 |
| Vigorous Q5 | Female | 1.02 | 1.00 | 1.03 | 0.07 |
| Moderate Q5 | Female | 1.00 | 0.98 | 1.01 | 0.61 |
| Walk Q5 | Female | 0.99 | 0.98 | 1.01 | 0.26 |
| Vigorous Q1 | Male | 1.01 | 0.98 | 1.03 | 0.55 |
| Moderate Q1 | Male | 0.99 | 0.98 | 1.01 | 0.43 |
| Walk Q1 | Male | 1.00 | 0.99 | 1.02 | 0.79 |
| Vigorous Q2 | Male | 1.01 | 0.99 | 1.03 | 0.38 |
| Moderate Q2 | Male | 1.02 | 1.00 | 1.04 | 0.11 |
| Walk Q2 | Male | 0.99 | 0.97 | 1.00 | 0.06 |
| Vigorous Q3 | Male | 1.01 | 0.99 | 1.02 | 0.29 |
| Moderate Q3 | Male | 1.00 | 0.98 | 1.02 | 0.92 |
| Walk Q3 | Male | 0.99 | 0.98 | 1.01 | 0.27 |
| Vigorous Q4 | Male | 1.05 | 1.03 | 1.07 | <0.001 |
| Moderate Q4 | Male | 1.00 | 0.98 | 1.02 | 0.99 |
| Walk Q4 | Male | 0.96 | 0.94 | 0.97 | <0.001 |
| Vigorous Q5 | Male | 1.06 | 1.04 | 1.08 | <0.001 |
| Moderate Q5 | Male | 0.98 | 0.96 | 1.00 | 0.06 |
| Walk Q5 | Male | 0.95 | 0.93 | 0.97 | <0.001 |

OR = Odds Ratio, CI = Confidence Interval

*Supplementary Table S11: Adjusted odds ratios for WHO physical activity categories and self-reported fracture, stratified by sex*

| Sex | Model | Category | Odds ratio | CI lower | CI upper | P value | Global P value |
|---|---|---|---|---|---|---|---|
| Male | Unadjusted | Meets guidelines | 1.02 | 0.95 | 1.09 | 0.596 | <0.001 |
| Male | Unadjusted | Exceeds guidelines | 1.32 | 1.26 | 1.39 | <0.001 | NA |
| Male | Minimal adjustment | Meets guidelines | 1.02 | 0.96 | 1.09 | 0.511 | <0.001 |
| Male | Minimal adjustment | Exceeds guidelines | 1.32 | 1.26 | 1.39 | <0.001 | NA |
| Male | Fully adjusted | Meets guidelines | 1.02 | 0.96 | 1.09 | 0.507 | <0.001 |
| Male | Fully adjusted | Exceeds guidelines | 1.31 | 1.24 | 1.38 | <0.001 | NA |
| Female | Unadjusted | Meets guidelines | 0.96 | 0.91 | 1.02 | 0.173 | <0.001 |
| Female | Unadjusted | Exceeds guidelines | 1.06 | 1.02 | 1.11 | 0.008 | NA |
| Female | Minimal adjustment | Meets guidelines | 0.96 | 0.91 | 1.02 | 0.194 | <0.001 |
| Female | Minimal adjustment | Exceeds guidelines | 1.07 | 1.02 | 1.11 | 0.006 | NA |
| Female | Fully adjusted | Meets guidelines | 0.96 | 0.91 | 1.02 | 0.185 | <0.001 |
| Female | Fully adjusted | Exceeds guidelines | 1.06 | 1.01 | 1.11 | 0.012 | NA |

OR = Odds Ratio, CI = Confidence Interval

### Supplementary Figures

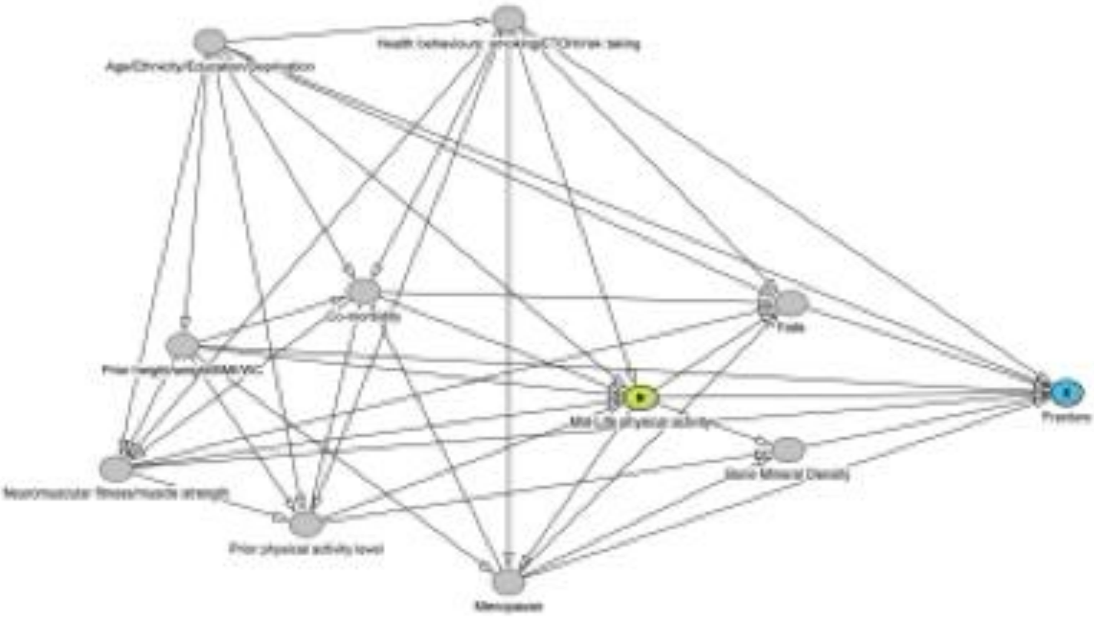


***Supplementary Figure S1***:*Directed Acyclic Graph of the possible pathways from mid-life physical activity to fracture. The directed acyclic graph (DAG) summarises the hypothesised relationships between mid-life physical activity (exposure), fracture (outcome), and measured covariates. Arrows represent assumed causal relationships based on prior evidence. Age, ethnicity, education/deprivation, health behaviours, prior physical activity, body size, neuromuscular fitness, comorbidity, and menopause were considered potential confounders. Falls and bone mineral density were considered potential mediators of the association between physical activity and fracture and were not included in the primary adjustment set. The green circle with a triangle denotes the exposure, and the blue circle the outcome.*

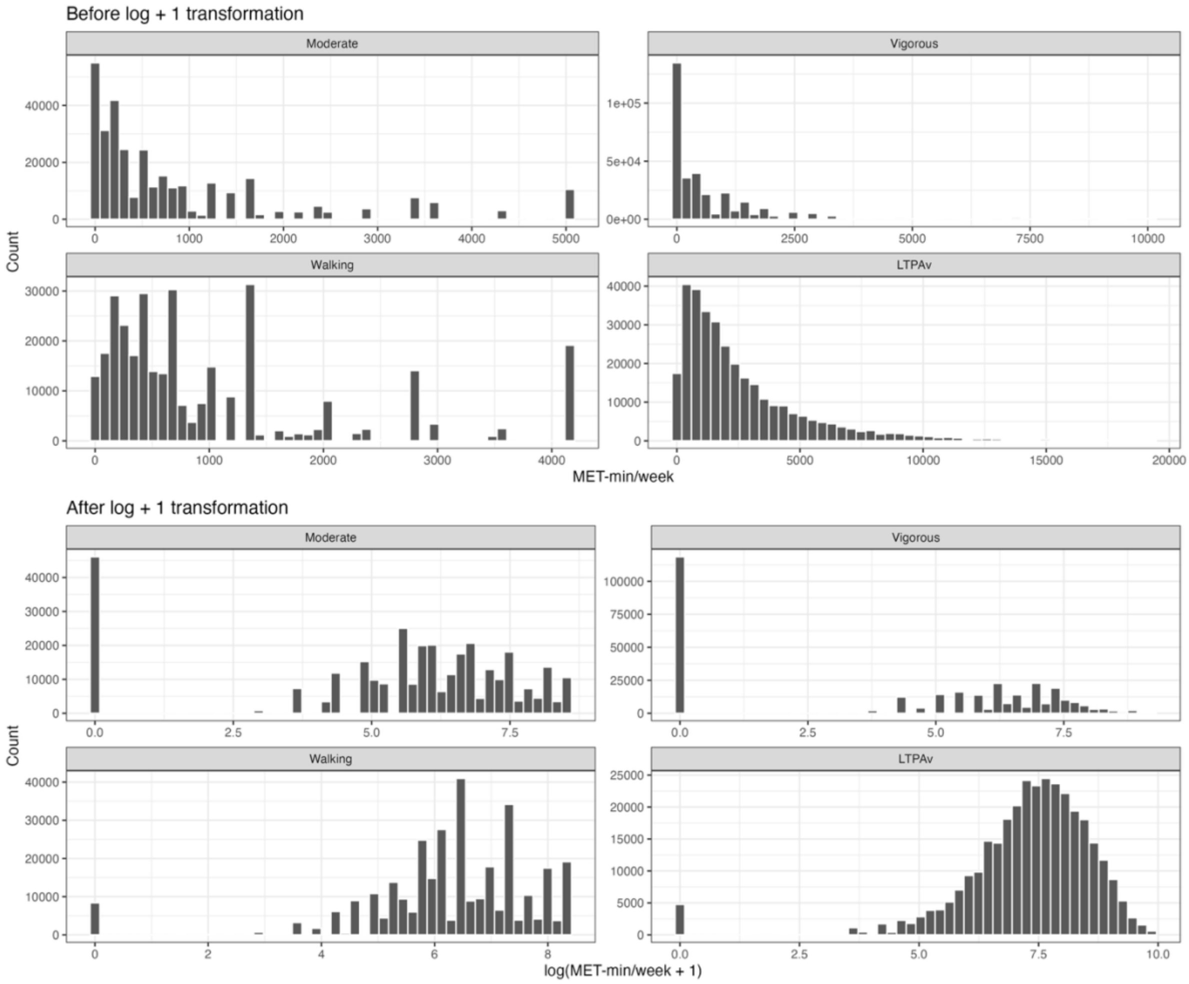


***Supplementary Figure S2.*** *Distribution of physical activity variables before and after log(MET-min/week + 1) transformation****.*** *Histograms show the distributions of moderate-intensity activity, vigorous-intensity activity, walking, and* $LTPA_V$ *(MET-min/week) before (top panels) and after (bottom panels) log(MET-min/week + 1) transformation. The transformation reduced the strong positive skew observed in the original distributions and improved their suitability for modelling as continuous exposures in restricted cubic spline analyses.*

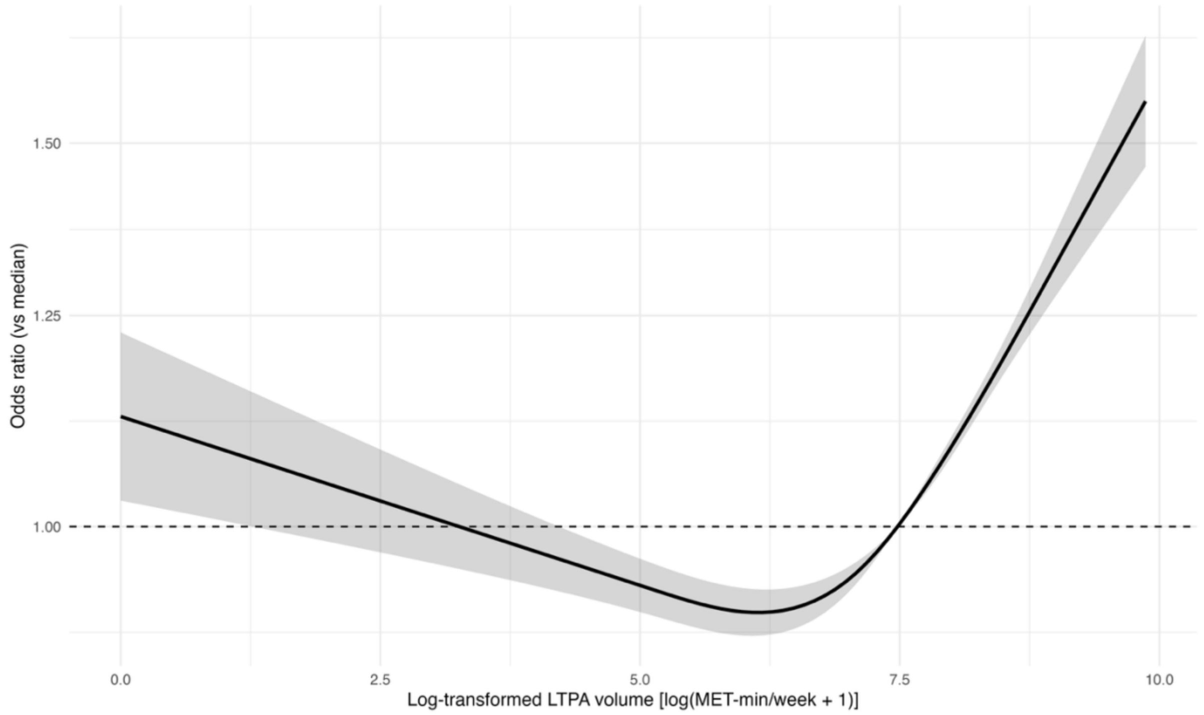


***Supplementary Figure S3. Association between total physical activity and fracture risk.*** *Association between log-transformed LTPA$_V$ (log[MET-min/week + 1]) and fracture risk estimated using a restricted cubic spline model. The solid line represents the adjusted odds ratio relative to the median physical activity level, and the shaded area indicates the 95% confidence interval. The dashed horizontal line represents an odds ratio of 1.0 (reference).*

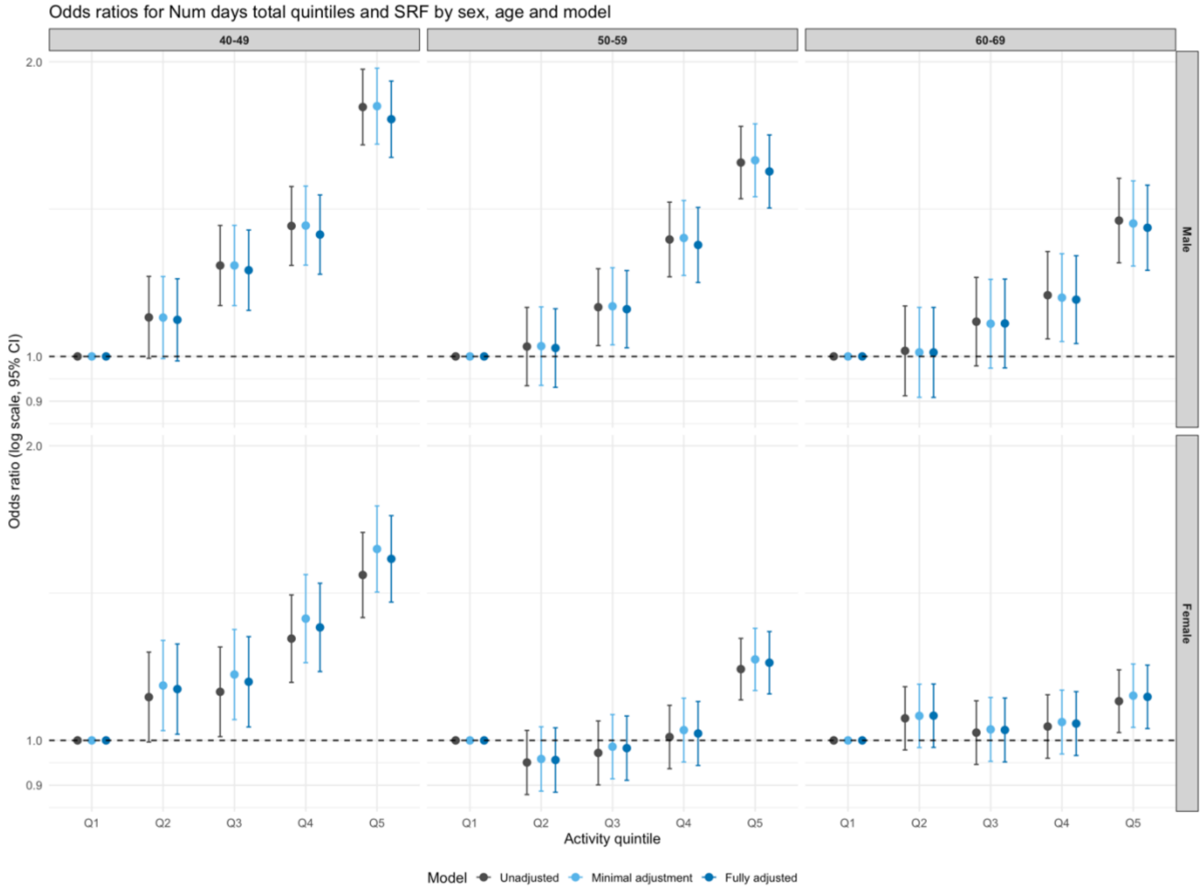


***Supplementary Figure S4:*** *Sensitivity analysis of the association between total physical activity frequency (days/week) and self-reported fracture by sex and age group. Odds ratios (95% confidence intervals) for self-reported fracture (SRF) according to quintiles of frequency of LTPA (days/week), stratified by sex and age group (40–49, 50–59, and 60–69 years). Estimates are shown for unadjusted, minimally adjusted, and fully adjusted logistic regression models. Quintile 1 (lowest activity frequency) was used as the reference category. This analysis was conducted as a sensitivity analysis using frequency of activity (days/week) rather than* $LTPA_V$ *(MET-min/week).*

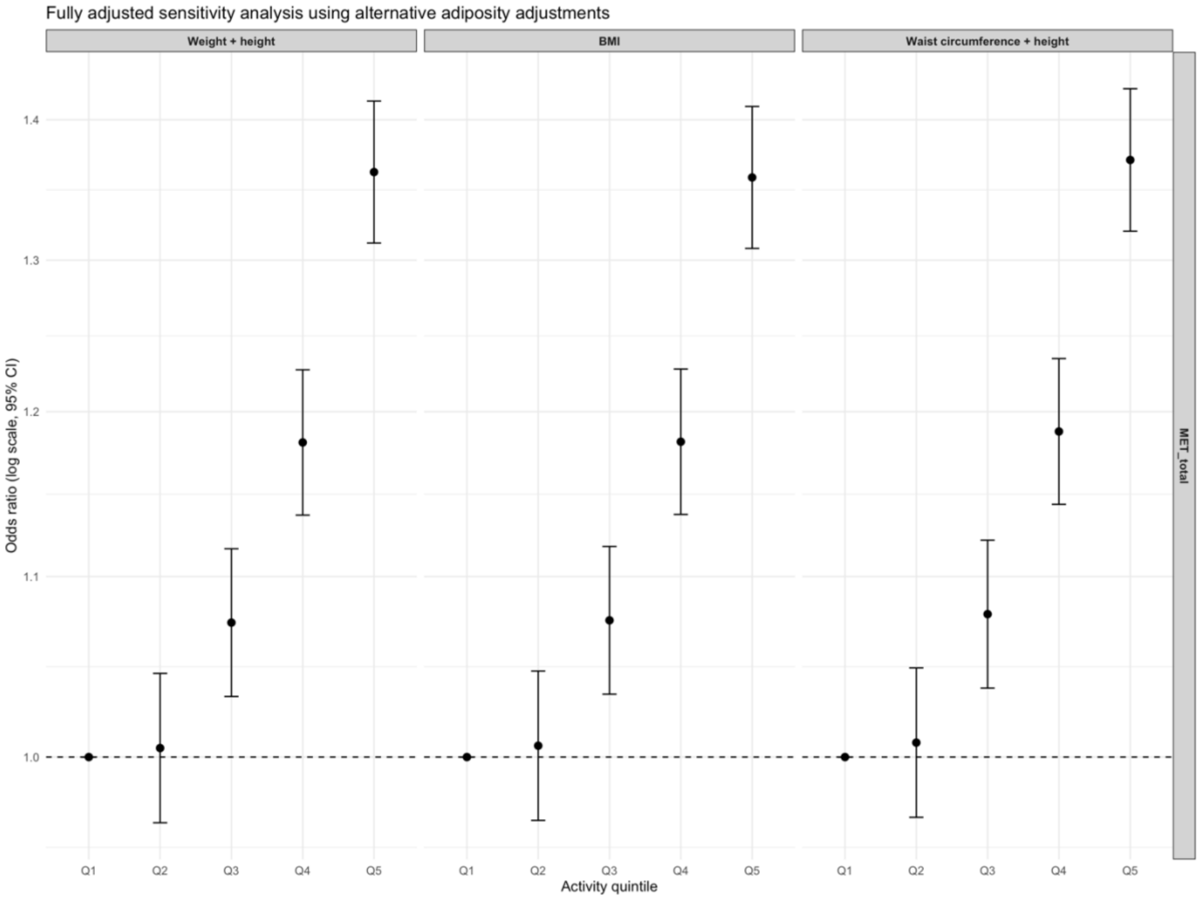


***Supplementary Figure S5.*** *Odds ratios (95% confidence intervals) for self-reported fracture (SRF) across quintiles of* $LTPA_V$ *from fully adjusted logistic regression models using alternative adiposity adjustment specifications: weight and height, body mass index (BMI), or waist circumference and height. Quintile 1 (lowest physical activity) was used as the reference category. Results were consistent across adiposity adjustment specifications, indicating that the observed associations were robust to the choice of adiposity measure.*

.

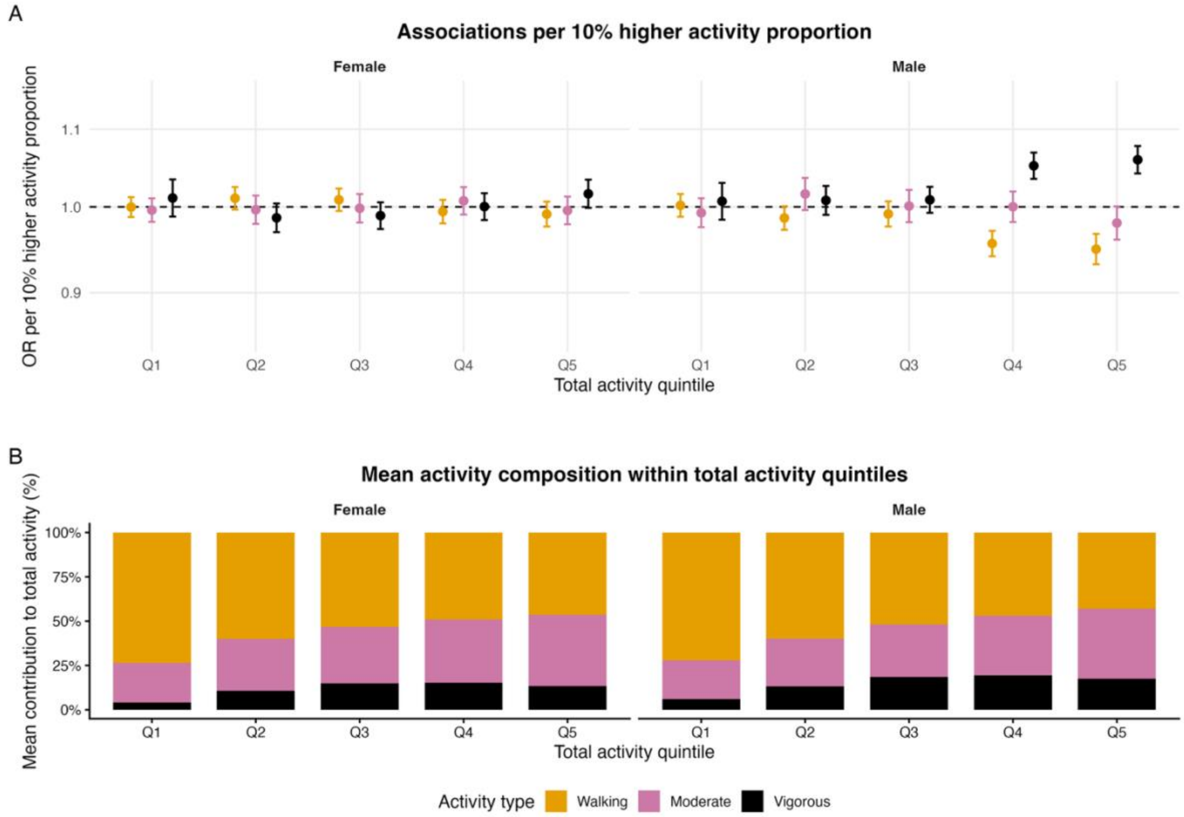


***Supplementary Figure S6.*** *(A) Odds ratios (ORs) and 95% confidence intervals for fracture associated with a 10% higher proportion of walking, moderate-intensity, or vigorous physical activity, while holding $LTPA_V$ constant, stratified by sex and total physical activity quintile. (B) Mean proportion of $LTPA_V$ contributed by walking, moderate-intensity, and vigorous activity within each $LTPA_V$ quintile, stratified by sex.*
*Total activity quintile = $LTPA_V$*
